# Epigenetic attenuation of interferon signaling drives aging-related improvements in systemic lupus

**DOI:** 10.1101/2025.01.27.25321143

**Authors:** Rithwik Narendra, Hoang Van Phan, Sarah L. Patterson, Ana Laura Almonte Loya, Cristina Lanata, Christina Love, Joonsuk Park, Emily C. Lydon, Michiko Ametani Shimoda, Lisa Barcellos, Honey Mekonen, Angela Detweiler, Padmini Deosthale, Norma Neff, Lindsey A. Criswell, Lenka Maliskova, Walter Eckalbar, Gabriella Fragiadakis, Jinoos Yazdany, Maria Dall’Era, Patricia Katz, Chun Jimmie Ye, Marina Sirota, Charles R. Langelier

**Affiliations:** Division of Infectious Diseases, University of California, San Francisco, CA, USA; University of California, Los Angeles, CA, USA; Division of Rheumatology, University of California, San Francisco, CA, USA; Department of Epidemiology and Biostatistics, San Francisco, CA, USA; National Human Genome Research Institute, Bethesda, Maryland, USA; Department of Experimental Medicine, University of California, San Francisco, CA, USA; University of California, Berkeley, CA, USA; Chan Zuckerberg Biohub San Francisco, San Francisco, CA, USA; UCSF Co-Labs, University of California, San Francisco, CA, USA; Bakar Computational Health Sciences Institute, University of California, San Francisco, CA, USA

## Abstract

In the general human population, aging is associated with a rise in systemic inflammation, primarily involving innate immune pathways related to interferon (IFN), toll-like receptor, and cytokine signaling. In systemic lupus erythematosus (SLE), a prototypical systemic autoimmune disease, aging is distinctly associated with improvements in disease activity, suggesting a unique relationship between aging and inflammation in this disease. Using a multi-omic approach incorporating transcriptional profiling, single cell RNA sequencing, proteomics and methylation analysis, we studied age-related changes in the immune profiles of 287 SLE patients between 20 and 83 years old, and compared the results against 928 healthy controls aged between 21 and 89 years old. In contrast to the increase in inflammatory gene expression that occurs with aging in most healthy adults, SLE patients exhibited the opposite. Most notable was a decrease in type I IFN signaling that was evident across multiple cell types, with CD56-dim natural killer (NK) cells, CD4^+^ effector memory T cells, and naïve B cells exhibiting the most significant differences. We found that aging in SLE patients was also associated with decreased IFN-α2 and IFN-λ1 levels, and differential methylation of the genome. Notably, of the genes both downregulated and hypermethylated with older age, IFN-related genes were disproportionately represented, suggesting that age-related decreases in IFN signaling were driven in part by epigenetic silencing. Both SLE patients and healthy controls demonstrated age-related declines in naïve T cells and lymphoid progenitor cells, but only SLE patients demonstrated age-related increases in CD56-dim NK cells. Taken together, our work provides new insight into the phenomenon of inflammaging and the unique clinical improvement in disease activity that occurs in SLE patients as they age.

## Introduction

Aging in the healthy human population is accompanied by an increase in systemic inflammation, predominantly involving innate immune pathways related to interferon (IFN), toll-like receptor, and cytokine signaling^1–4^. This state of age-related immune dysregulation, commonly referred to as inflammaging, is implicated in the pathophysiology of several major chronic diseases such as cardiovascular disease, diabetes, cancer, and osteoarthritis, each of which typically increases in both prevalence and severity as one ages^5^. While the mechanisms underlying age-related inflammation are incompletely understood, a growing body of literature implicates mitochondrial dysfunction, oxidative stress, and the accumulation of damage-associated molecular patterns (DAMPs), which together drive inflammatory gene expression and cytokine release^6^.

Systemic lupus erythematosus (SLE) is a prototypical autoimmune disease characterized by loss of tolerance to nuclear antigens, autoantibody production, and pathogenic inflammation involving multiple organ systems. Affected individuals may experience debilitating symptoms, impaired quality of life, and premature mortality^7^. Interestingly, in contrast to most chronic inflammatory diseases, systemic inflammation from SLE appears to improve in older age. For instance, late-onset SLE is characterized by lower disease severity compared to patients with disease onset under 50 years of age^8,9^, and increasing age is associated with both lower disease activity^9–11^ and lower risk of developing lupus nephritis^12^. Though age-related improvements in SLE are commonly observed in clinical practice, the mechanisms for this unique inverse relationship between age and systemic inflammation in SLE are poorly understood.

Here, we seek to examine the relationship between age and pathogenic inflammation in SLE by conducting multiomic analysis—whole blood transcriptional profiling, single-cell RNA sequencing (scRNA-seq) and epigenetic analysis—on individuals with SLE across the adult age spectrum. We find a distinct reversal of age-related innate immune activation, most notably involving IFN signaling, in patients with SLE, and discover that this relationship is partly mediated by epigenetic changes in type-I IFN genes.

## Results

### Study cohorts

We performed a prospective observational study of 287 SLE patients enrolled in the California Lupus Epidemiology Study (CLUES)^13^. SLE participants ranged in age from 20–83 years, were predominantly female, and represented diverse racial and ethnic backgrounds (**Fig. 1, Supp. Fig. 1, Supp. Tables 1A, 1B**). We evaluated the impact of aging on gene expression, biological pathway activation, immune cell populations, protein expression and DNA methylation using a combination of whole blood bulk RNA-seq (n=271), peripheral blood mononuclear cell (PBMC) scRNA-seq (n=148), proximity extension assay proteomics (n=268), and bisulfite sequencing (n=267) (**Fig. 1, Supp. Fig. 2A-C**). We next compared transcriptomic findings in SLE patients against those from healthy controls ranging in age from 21–89 years, leveraging data from the Rotterdam population surveillance study^14^ (n=880, whole blood microarray, **Supp. Data 1**) and the CLUES cohort (n=48, scRNA-seq, **Supp. Data 2**).

**Figure 1.**
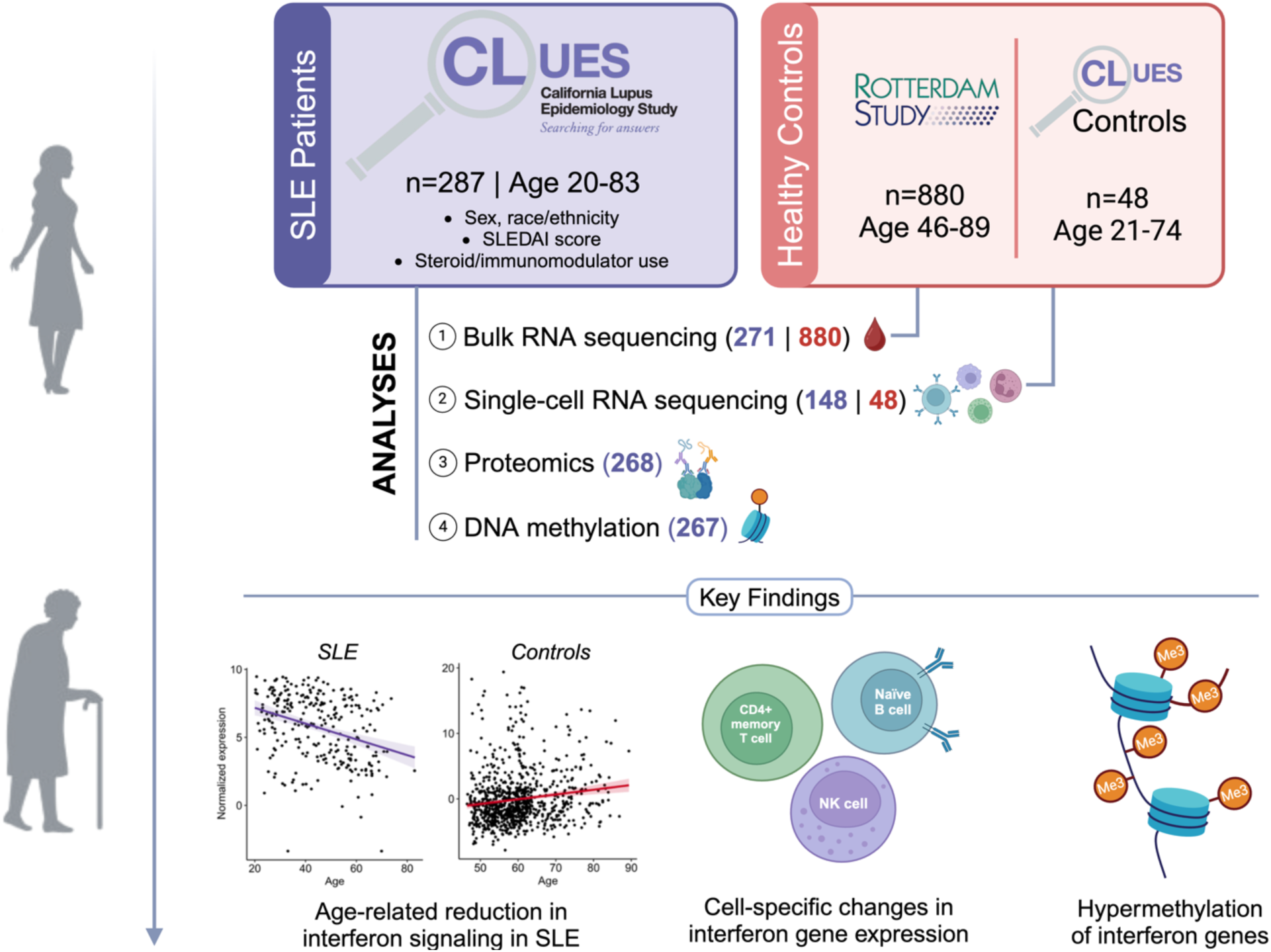
Study Overview. This study evaluated 287 patients with systemic lupus erythematosus (SLE) and 928 healthy controls. Age related differences in gene expression, biological pathway activation and immune cell populations were assessed in blood using bulk RNA sequencing (n=271 SLE, n=880 healthy controls) and single-cell RNA sequencing (scRNA-seq) (n=148 SLE, n=48 healthy controls). Epigenetic changes were assessed with bisulfite sequencing (n=267 SLE).

### SLE disease activity decreases with age

We first asked whether age was associated with changes in SLE disease activity within the CLUES cohort by calculating the Systemic Lupus Erythematosus Disease Activity Index (SLEDAI) score^15^ for study participants as a function of age. We found a significant negative relationship between age and SLEDAI score after controlling for sex and race/ethnicity (P = 1.5e-3). This demonstrated that, as expected, SLE disease activity improved with higher age in our cohort (**Supp. Fig. 3**).

### Aging in SLE patients is associated with downregulation of interferon and other innate-immune response genes

We next analyzed whole blood RNA-seq data from SLE patients (n=271) to evaluate age-related changes in gene expression. We identified 319 genes significantly associated with age (adjusted P (P_adj_) < 0.05), controlling for sex and race/ethnicity, and treating age as a continuous variable (**Fig. 2A, Supp. Data 3**). Using gene set enrichment analysis (GSEA), we explored the biological function of these genes and found that older age was associated with a striking downregulation of innate immune pathways, most notably IFN-α/β and IFN-γ signaling (**Fig. 2B, Supp. Data 3**). Repeating this analysis in exclusively female SLE patients (n=240) yielded 300 genes significantly associated with age and the same significantly downregulated pathways on GSEA (**Supp. Fig. 4, Supp. Data 4**).

**Figure 2.**
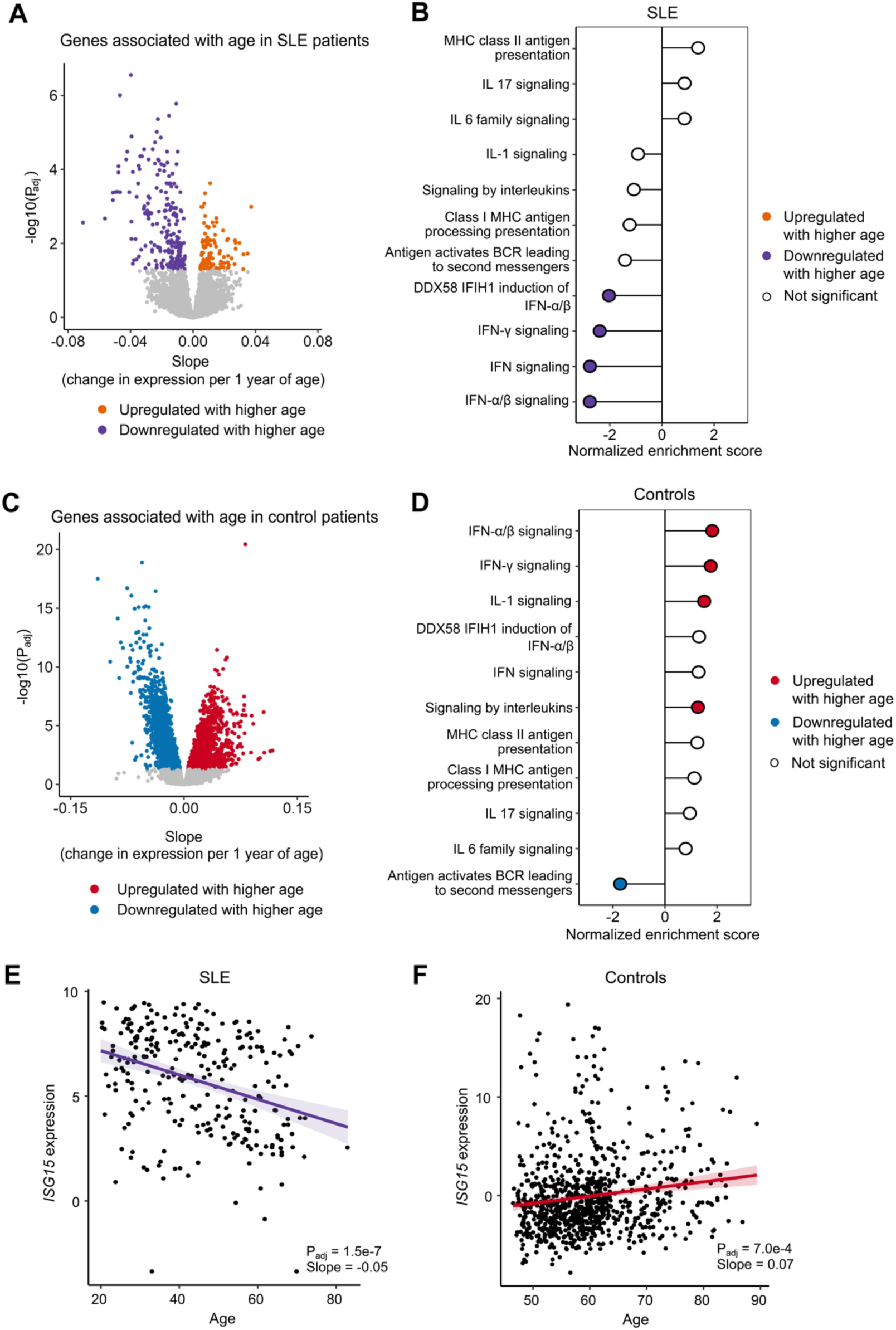
Aging associates with reduced interferon gene expression in SLE patients but not healthy controls. **A** Volcano plot highlighting genes with significant age-related changes in expression in SLE patients (n=271), treating age as a continuous variable. P values were calculated using the limma package and adjusted with the Benjamini-Hochberg method. **B** Gene Set Enrichment Analysis (GSEA) demonstrating immune-related biological signaling pathways in SLE patients. Filled circles represent pathways with a Benjamini-Hochberg adjusted P value < 0.05. A positive normalized enrichment score value represents upregulation of the pathway with older age and a negative value represents downregulation with older age. **C** Volcano plot highlighting genes with significant age-related changes in expression in healthy controls (n=880), treating age as a continuous variable. P values were calculated using the limma package and adjusted with the Benjamini-Hochberg method. **D** GSEA demonstrating immune-related biological signaling pathways in healthy controls. **E, F** Linear regression assessing relationship between expression of the canonical interferon stimulated gene *ISG15* and age in (E) SLE patients and (F) healthy controls. Expression was measured by log2 counts per million. Slope refers to the change in normalized gene expression per year of age. P values were calculated using limmaand adjusted with the Benjamini-Hochberg method.

Additionally, to investigate whether differences in treatment with immunosuppressive agents might have influenced our results, we performed a sensitivity analysis adjusting for treatment with systemic glucocorticoids among the SLE patients for whom we had medication information (n=267). Our results remained largely unchanged, with GSEA still demonstrating significant downregulation of (IFN)-α/β, IFN-γ and other innate immune-related pathways (**Supp. Fig. 5, Supp. Data 5**). We also performed sensitivity analyses adjusting for hydroxychloroquine treatment (**Supp. Fig. 6, Supp. Data 6**) or body mass index **(Supp Fig. 7, Supp. Data 7**), and found that in both analyses results were largely unchanged.

Next, we sought to perform a similar analysis in healthy control patients by leveraging a large public blood transcriptomic dataset from the Rotterdam Study^14^ (n=880). Healthy control participants from this study ranged from 46-89 years old, 46% of whom were women. Analyzing all individuals, we identified 3972 genes significantly associated with age, controlling for sex (P_adj_ < 0.05, **Fig. 2C, Supp. Data 8**). Consistent with prior studies of aging and blood gene expression^1–4^, GSEA demonstrated broad upregulation of innate immune signaling pathways with older age, including many of the same pathways that were downregulated with age in SLE participants (**Fig. 2D**). In addition, we observed significant downregulation of B-cell receptor activation pathways with age, consistent with prior observations demonstrating impaired adaptive immune responses in older adults^3,16^.

The opposing directionality of age-related changes in canonical interferon-stimulated gene (ISG) expression between SLE participants and healthy controls was well exemplified by *ISG15*, the expression of which significantly decreased with age in SLE patients (P_adj_ = 1.5e-7, **Fig. 2E**) but increased with age in the healthy controls (P_adj_ = 7.0e-4, **Fig. 2F**). We noted that some ISGs were statistically significantly downregulated with higher age in SLE patients (e.g., *IFIT1*) without significant age-related changes in controls, while others are both significantly downregulated with higher age in SLE and significantly upregulated with age in controls (e.g., *IFI27*) (**Supp. Fig. 8**). We calculated an IFN-α/β score based on the average expression of genes in the IFN-α/β signaling pathway, and found that it significantly decreased with older age in SLE patients (**Supp. Fig. 9A**) but demonstrated the opposite relationship in controls (**Supp. Fig. 9B**). We observed a similar dynamic for an IFN-γ score (**Supp. Figs. 9C, D**). Adjusting for age of SLE diagnosis did not significantly alter the inverse relationship between age of SLE patient and either of the calculated IFN scores (**Supp. Fig. 10A, B**). Collectively, these findings point to decreasing ISG expression as an important mediator for clinical improvements in disease activity observed among individuals with SLE as they grow older.

### Aging in SLE patients is associated with interferon downregulation at the protein level

We next investigated whether these observations could be generalized to the protein level by measuring the plasma concentrations of 48 inflammatory cytokines (**Supp. Data 9**) in SLE patients using the Olink proteomics proximity extension assay. We found that levels of 12 proteins were significantly associated with age (**Fig. 3A, Supp. Data 9**). Notably, we found that IFN-α2 (**Fig. 3B**) and IFN-λ1 were significantly downregulated with higher age, demonstrating that age-related decreases in interferon signaling occur both at the transcriptional and protein levels in SLE. We also recognized that several of the upregulated proteins (e.g., TREM1, FGF21) have previously been associated with older age and inflammaging^17,18^.

**Figure 3.**
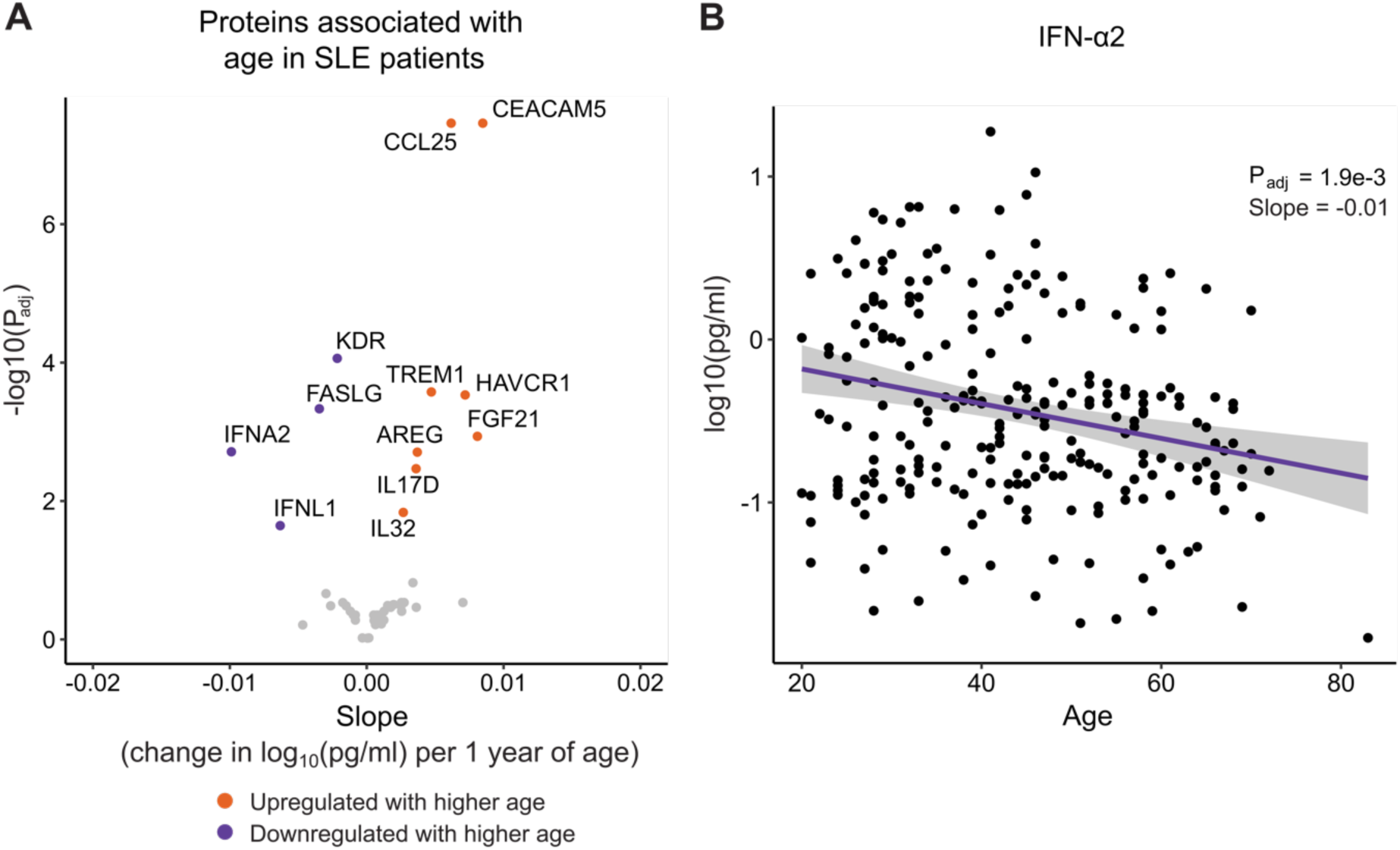
Proteomic analysis demonstrates that aging associates with lower interferon levels in SLE patients. **A.** Volcano plot highlighting inflammatory proteins significantly associated with age in SLE patients (n=268), treating age as a continuous variable. 48 inflammatory proteins measured by Olink proximity extension assay. All concentrations were log10-transformed after the addition of a small constant (10^−6^) to avoid taking the log of 0. The ribbon represents the confidence interval along the regression line. **B.** Linear regression assessing the relationship between expression of IFN-α2 and age in SLE patients. Slope refers to the change in protein concentration per year of age. In A and B, P values were calculated using linear regression and adjusted with the Benjamini-Hochberg method.

### Age-dependent changes in immune cell populations differs in SLE patients compared to healthy controls

Next, we sought to evaluate age-related transcriptional changes at the single cell level in PBMCs from SLE patients (n=148) and healthy controls from the CLUES cohort (n=48) (**Supp. Table 1B**). Leveraging scRNA-seq data^19^ from 1.2 million PBMCs across all patients, we identified a diversity of immune cell populations (**Fig. 4A, Supp. Fig. 11**). The proportions of several cell types varied with age, with differences observed based on SLE status (**Fig. 4B**). For instance, proportions of CD56-dim NK (NK_dim_) cells, a NK cell subpopulation with greater cytotoxic activity^20^, demonstrated a significant positive association with age in the SLE group, a trend that was not observed in healthy controls. Conversely, only controls demonstrated age-dependent increases in the proportions of CD4+ effector memory T cells (CD4^+^ T_EM_) and CD4+ regulatory T cells (CD4^+^ T_reg_). In both SLE and healthy control patients, the proportions of naïve CD8+ T cells (CD8^+^ T_naïve_) and progenitor cells (Progen) declined with age (**Fig. 4B**).

**Figure 4.**
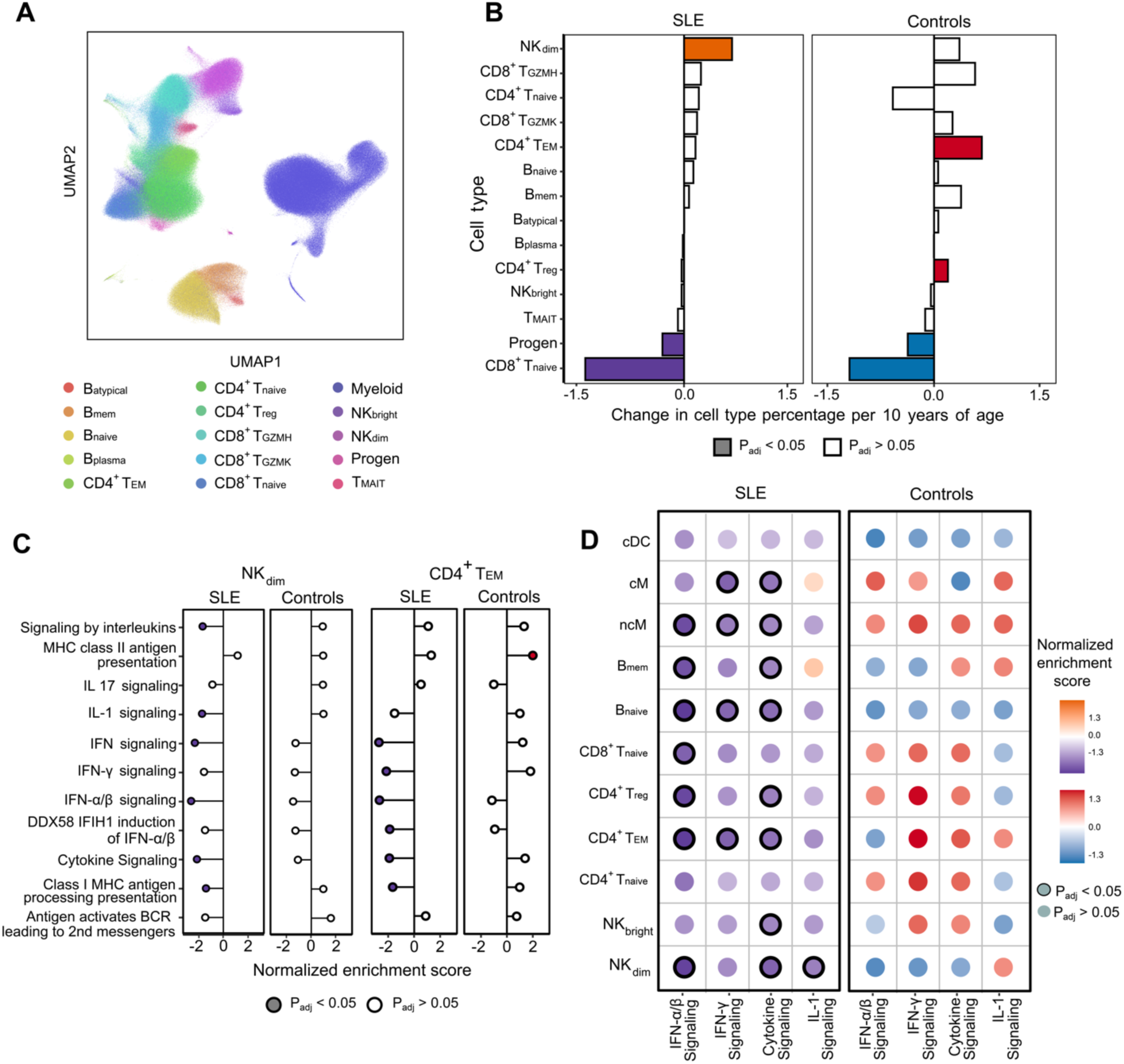
Aging associates with different immune cell frequencies and reduced expression of innate immunity genes in SLE patients versus controls. **A** Uniform Manifold Approximation and Projection plot of immune cell types identified by scRNA-seq (n=148 SLE patients, n=48 healthy controls). **B** Bar plots depicting change in the percentages of immune-related blood cell types with age, including those that are significantly increased (orange for SLE, red for controls) or decreased (purple for SLE, blue for controls). Statistical significance (indicated by filled bars) determined based on an adjusted P value < 0.05, calculated using linear modeling with Benjamini-Hochberg correction. **C** Gene Set Enrichment Analysis (GSEA) demonstrating changes in immune-related signaling pathways from the Reactome database, in SLE patients and in healthy controls for NK_dim_ and CD4^+^ T_EM_ cells. A positive normalized enrichment score (NES) represents upregulation of the pathway with older age, and a negative value represents downregulation with older age. Filled circles represent significant pathways with a Benjamini-Hochberg adjusted P value < 0.05. **D** Dot plots depicting the NES of four immune-related signaling pathways from panel C for each cell type, for SLE patients (left) or healthy controls (right). For each group, upregulation (orange for SLE and red for controls) or downregulation (purple for SLE and blue for controls) of pathway is determined based on the NES. Black outlines on dots indicate significant (based on Benjamini-Hochberg adjusted P value < 0.05) regulation. cDC, conventional dendritic cell; cM, classical monocyte; ncM, non-classical monocyte.

### Age-dependent immune cell gene expression differs between SLE patients and healthy controls

To determine which immune cell populations were driving age-related changes in innate immune signaling in SLE patients, we performed pseudobulk differential gene expression analysis within each cell type, treating age as a continuous variable and adjusting for sex and race/ethnicity. In SLE patients, we found that several cell types exhibited age-related changes in gene expression (**Supp. Fig. 12**). Amongst lymphocytes, these included the NK_dim_ cells, naïve B cells (B_naïve_), CD8^+^ T_naïve_, CD4^+^ T_EM_, CD4^+^ T_naïve_ and CD4^+^ T_reg_. NK_dim_, CD4^+^ T_EM_ and B_naïve_ cells exhibited the greatest age-associated changes in gene expression (**Figs. 4C, D**), reflecting significant downregulation of many of the key pathways found to be associated with age in the bulk RNA-seq analysis (**Fig. 2C**), including IFN-α/β, IFN-γ, IL-1, and cytokine signaling. This suggested that transcriptional changes in NK_dim_ and CD4^+^ T_EM_ cells with respect to age may be drivers of this trend. While GSEA of scRNA-seq data from a comparatively small cohort of healthy control patients did not yield many significantly enriched pathways (**Fig. 4C, D**), genes related to IFN-α/β and IFN-γ signaling were upregulated with age in most T-cell and monocyte populations (**Fig. 4D**).

### Aging leads to hypermethylation of interferon-related genes in SLE patients

Hypothesizing that epigenetic modification might underlie the age-dependent downregulation of innate immune gene expression observed in SLE, we evaluated the DNA methylation status of CpG sites from 267 patients in the CLUES cohort. We found 48,675 CpGs hypermethylated with age and 61,980 hypomethylated with age out of a total 646,554 CpGs (**Fig. 5A**). Of the 2,035 CpGs mapped to genes with age-related decreases in expression by bulk RNA-seq analysis, 230 were hypermethylated (P=5.3e-10 by hypergeometric test, **Fig. 5B**). Intriguingly, within this set of 230 CpGs, the majority (n=133) mapped to genes that had biological functions related to IFN signaling (**Fig. 5C**). These included many of the same genes found to be most downregulated with age in SLE patients, and upregulated with age in healthy controls (**Fig. 2E, F**, **Fig. 5D**) as well as other canonical ISGs with roles in SLE pathogenesis (e.g., *RSAD2*^21,22^). As a complementary approach, we also assessed aging-associated genes for which methylation correlated with decreased expression, (**Supp. Fig. 13, Supp. Data 10**), and noted that most of the significantly correlated genes (91%) were related to IFN signaling. Together, these findings suggested that in SLE, age-related hypermethylation of IFN-related genes could contribute to their suppressed expression.

**Figure 5.**
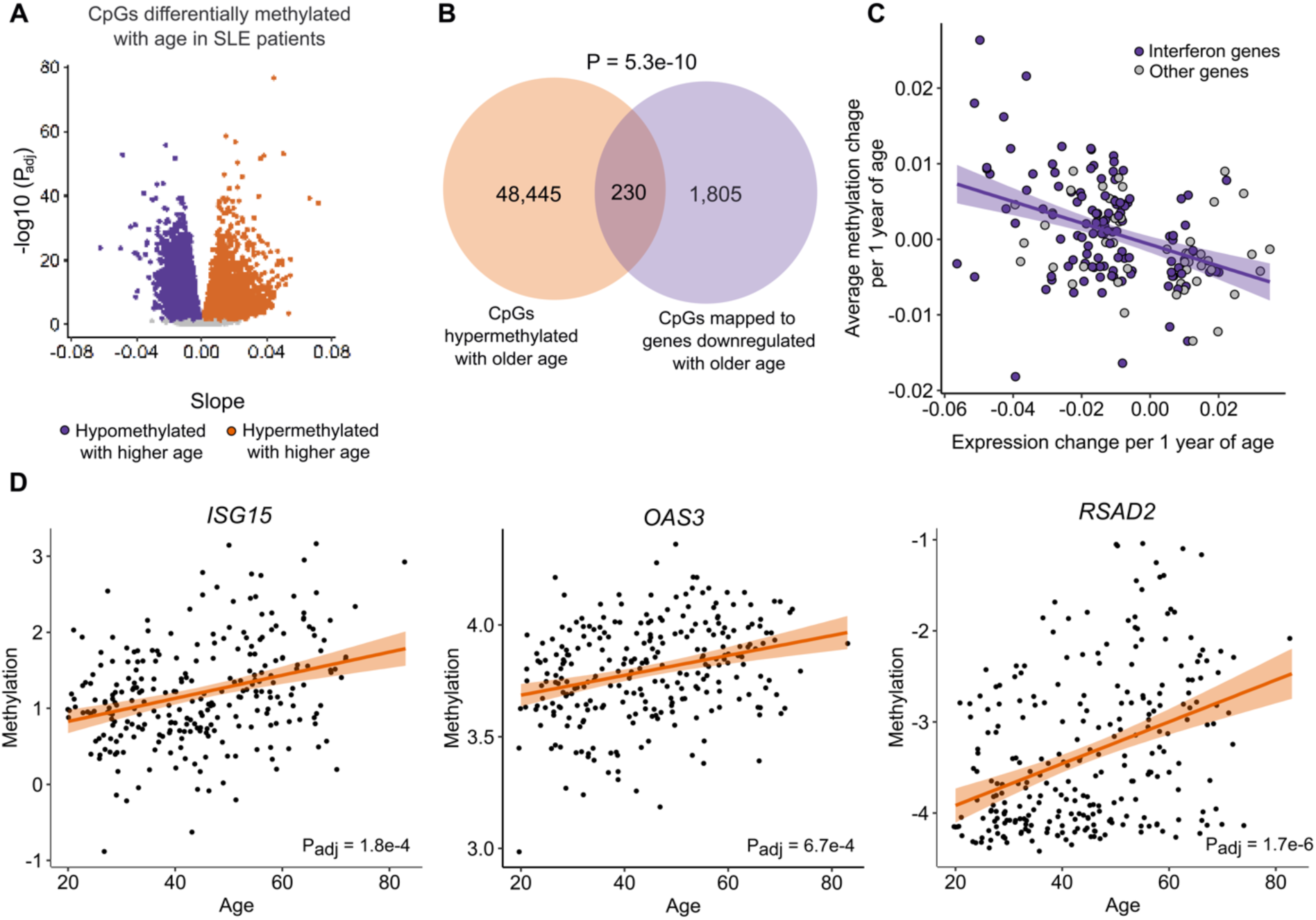
Aging results in hypermethylation of interferon-related genes in SLE. **A** Volcano plot demonstrating differentially methylated CpGs based on age at an adjusted P value < 0.05. **B** Hypermethylated CpGs mapping to genes significantly downregulated with age. P value calculated based on hypergeometric test. **C** Scatter plot showing the relationship between age-associated changes in gene expression by bulk RNA-seq and changes in CpG methylation levels in SLE patients, focusing on the 230 overlapping CpGs from panel B. CpGs mapping to interferon-related genes (n=133) are highlighted in purple, the remaining genes are plotted in gray. **D** Age-related changes in methylation levels of CpGs mapping to representative interferon-related genes in SLE patients, including *ISG15* (cg04788999), *OAS3* (cg08147692), and *RSAD2* (cg10771443). The lines indicate the general relationship between age and methylation for the gene, and the ribbons indicate the confidence interval for this estimate. P values were calculated using linear modeling and were adjusted with the Bonferroni-Hochberg method.

### Innate immune signaling remains persistently higher in SLE patients versus controls across the age spectrum

Our results so far demonstrated opposing directionality in age-related changes in innate immune gene expression between SLE patients and controls. Whether these trajectory differences ultimately resulted in complete normalization of inflammatory signaling in SLE patients in older age remained unclear. We thus asked how innate immune profiles compared between SLE patients and controls within three different age groups (under 50, over 50, and over 65 years old), using our scRNA-seq data. We restricted this analysis to females, to avoid confounding by known sex-based differences in aging-related gene expression.

We identified 5,435 genes significantly associated with SLE status in female patients under 50 years of age (n=110), controlling for race/ethnicity (P_adj_ < 0.05, **Supp. Fig. 14A**). GSEA demonstrated significant upregulation of type I and type II IFN signaling in SLE patients compared to controls (**Supp. Fig. 14B**). Repeating these analyses in patients over 50 years of age (n=71) demonstrated similar upregulation of innate immune pathways in SLE **(Supp. Fig. 14C, D**), as did the same analysis in patients over 65 years of age (n=17) **(Supp. Fig. 14E, F).** Collectively, these findings demonstrated that while there is a reduction in IFN signaling with older age in SLE patients, innate immune signaling nevertheless remains significantly upregulated in SLE patients compared to healthy controls across the age spectrum.

## Discussion

In this study, we report that SLE patients demonstrate a unique relationship between increasing age and inflammatory gene expression compared to the general population. As opposed to an increase in the expression of innate immune genes with older age in healthy individuals, SLE patients exhibited the opposite, most notably for genes related to IFN signaling. Using scRNA-seq and methylation analysis, we demonstrate that this occurs across multiple cell types, and is mediated, at least in part, by epigenetic hypermethylation of IFN-related genes.

Across mammals, gradual increases in low level inflammation are an inevitable feature of older age, contributing to a number of chronic diseases that span a diverse range of organ systems^2,23,24^. Among the general population, a growing body of literature has demonstrated that increased activation of pattern recognition receptors with aging drives downstream increases in inflammatory gene and protein expression, including type I and II interferons^2,3,25^. This is juxtaposed against immunosenescence, characterized by diminished production of naïve T cells, impaired T cell signaling, and reduced responses of diverse immune cells to antigenic stimulation^26^. Our analyses of healthy control transcriptomic and scRNAseq data largely reflected these established findings.

In SLE, however, the relationship between age and systemic inflammation markedly differed compared to healthy controls. We initially considered the possibility that this could represent a complete reversal of inflammaging in SLE. However, a direct comparison of scRNA-seq data from female SLE patients and controls over 65 years of age demonstrated higher inflammatory gene expression in those with lupus. This finding suggests that while innate inflammatory pathways are downregulated with increasing age in SLE, expression of inflammatory genes remains elevated relative to healthy controls throughout the lifespan, even in older age.

Elevated IFN-α levels were first described in SLE patients more than 40 years ago^27^, and more recent studies have demonstrated that augmented type I IFN gene expression characterizes the blood transcriptome of most SLE patients^28,29^. Furthermore, higher IFN levels are associated with worse disease activity^30^, and directly contribute to the molecular pathogenesis of SLE^31,32^. While mechanisms underlying this remain incompletely understood, reduced ISG promoter methylation leading to augmented gene expression appears to be at least partially involved^33,34^.

Intriguingly, we find that in SLE, older age is associated with hypermethylation of multiple canonical ISGs such as *ISG15, OAS3* and *RSAD2*. Amongst genes both downregulated and hypermethylated with age in SLE patients, IFN-related genes are overrepresented. This suggests that age-related epigenetic modifications may drive suppression of type I IFN signaling and may at least partially explain the negative association between reduced SLE activity and advanced age observed in clinical practice^9,10,35^.

Aging is characterized by changes in immune cell populations^36^. In particular, adaptive immune responses decline with age, leading to a variety of complications ranging from diminished vaccine responses to less effective defense against microbial pathogens such as SARS-CoV-2^3,16^. Reflecting this, we found that both SLE patients and controls demonstrated age-related declines in naïve T cells and lymphoid progenitor cells. Only SLE patients, however, demonstrated age-related increases in NK_dim_ cells, which have greater cytotoxic potential and are the dominant NK cell subset in the peripheral blood^20^. NK cytopenia is a well-described feature of SLE^31,37^ that may be caused in part by elevated levels of IFN-α, which contributes to activation-induced apoptosis of NK cells^38^. It is thus possible that age-related decreases in IFN signaling contribute to the commensurate increases in NK_dim_ cells that we observed.

Of the immune cell populations studied, gene expression in NK_dim_, CD4^+^ T_EM_ and B_naïve_ cells was most significantly influenced by age in SLE patients. Notably, each of these cell types has been implicated in SLE pathogenesis. CD4^+^ T_EM_, for instance, are expanded in SLE^39^, and disruption of the relationship between CD4^+^ T_reg_ and CD4^+^ T_EM_ adversely impacts T cell activation and contributes to disease progression^40^. Given the importance of autoantibodies in SLE, B cells have a prominent role in disease pathogenesis and are the target of several important therapeutic agents^41^. Whether additional epigenetic modifications or alternative regulatory processes are responsible for age-related downregulation of interferon signaling at the gene and protein levels remains to be studied.

This study has several strengths, including affording new insights into the relationship between aging and inflammatory gene expression in SLE, a large sample size, detailed clinical phenotyping, and a multi-omic analytic approach incorporating bulk RNA-seq, scRNA-seq, proteomics and methylation assessment. Our study also has limitations, including a cross-sectional design versus a longitudinal design following individuals from youth into older age. Overall disease activity in the cohort was low, and future studies will be needed to determine if findings also extend to SLE patients with a greater range in SLEDAI scores. In our primary bulk transcriptomic analysis, we relied on publicly available microarray data to provide comparative transcriptomic assessment of healthy controls across the age spectrum, as we did not have healthy control patients with bulk RNA-seq data from the CLUES cohort. Fortunately, age-related changes in inflammatory gene expression have been extensively characterized, and the Rotterdam Study control group primarily served to recapitulate established results using a consistent analytic approach. Moreover, with scRNA-seq, we were able to directly compare inflammatory gene expression between SLE patients and controls using a single, consistent method. Finally, we also acknowledge the risk of survivorship bias as patients with severe lupus are at risk for premature mortality and thus may not have been well represented among the oldest patients in the CLUES cohort.

In summary, our findings indicate that patients with SLE exhibit age-related decreases in the expression of type I IFN and other innate immune genes in the peripheral blood, contrary to the trends observed in the general population.

## Methods

### Patient enrollment

We studied participants in the California Lupus Epidemiology Study (CLUES), an ongoing prospective longitudinal cohort of adults with SLE, as well as healthy controls. The UCSF Institutional Review Board approved this study (protocol number 14-14429) and informed consent was received prior to enrollment. SLE diagnoses were confirmed by study physicians based on (a) ≥4 of the 11 American College of Rheumatology (ACR) revised criteria for the classification of SLE^42,43^, (b) meeting 3 of the 11 ACR criteria with a documented rheumatologist’s diagnosis of SLE, or (c) a confirmed diagnosis of lupus nephritis. We evaluated CLUES participants from whom either PAXgene whole blood RNA tubes were collected or PBMC scRNA-seq data was available^19^.

All SLE patients were evaluated at a research clinic visit by a rheumatologist with expertise in SLE. In addition to blood collection, participants completed of a structured interview in which they were asked about sociodemographic characteristics, including sex, age, and race, smoking status and comorbidities including cardiovascular disease, diabetes mellitus, and cancer. Treatment with glucocorticoids or other immune modulating agents was also documented. SLE disease activity was measured using the Systemic Lupus Disease Activity Index (SLEDAI), also known as the SELENA-SLEDAI tool^44^.

### SLEDAI score versus age analysis

From all 271 samples in the CLUES bulk RNA-seq dataset, we fit a linear regression model for the SLEDAI score with the lm function using the following R design formula:

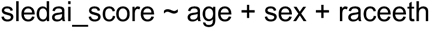

where sledai_score is the SLEDAI score of a given sample, age is the patient’s age, sex is a categorical variable with two levels (male or female), and raceeth is a categorical variable with four levels describing the race/ethnicity of the patient (White, Hispanic, Black, or Asian). For our regression plot, we extracted the slope and P value for age. We then created a confidence interval using the ggpredict function (terms = “age”) from the ggeffects package v1.3.2^45^.

### RNA extraction and sequencing

Following enrollment in CLUES, whole blood was collected in PAXgene tubes, processed according to manufacturer’s instructions and stored at -80°C. RNA was extracted using the Qiagen RNeasy kit and normalized to 20ng total input per sample. For RNA-seq library preparation, human cytosolic and mitochondrial ribosomal RNA and globin RNA was first depleted using FastSelect (Qiagen). RNA was then fragmented and underwent library preparation using the NEBNext Ultra II RNA-seq Kit (New England Biolabs) according to manufacturer’s instructions with protocol optimization for a LabCyte Echo acoustic liquid handler^46^. Finished libraries underwent 146 nucleotide paired-end Illumina sequencing on an Illumina Novaseq 6000 instrument.

### Bulk RNA-seq analysis of SLE patients from the CLUES cohort

All data analyses were done in R v4.3.1. For the SLE patient analyses, we evaluated differential gene expression using linear modeling with age as a continuous variable, and controlling for sex and race/ethnicity. For bulk RNA-seq analyses, we retained samples with at least 10,000 protein-coding genes. For the SLE patient bulk RNA-seq analyses, we retained protein-coding genes that had a minimum of 10 counts in at least 20% of the samples. We normalized the gene counts using the voom function (normalize.method = “quantile”) from the limma package v3.58.0^47,48^ and fitted a linear model for the gene expression with the lmFit function by using the following R design formula:

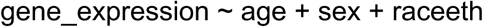

where gene_expression is the normalized expression of a given gene, age is the patient’s age, sex is the patient’s sex (one of two categories, male or female, defined as sex assigned at birth), and raceeth is the patient’s race/ethnicity (one of four categories: White, Hispanic, Black, or Asian). We then calculated the empirical Bayes statistics with eBayes function (default settings), and calculated the P values for differential expression with Benjamini-Hochberg multiple comparison correction.

We extracted the differential gene expression results using the topTable function (coef = “age”) from the limma package. We considered genes to be significantly differentially expressed if their adjusted P values were less than 0.05 (False Discovery Rate < 0.05). For our linear regression gene plots, we normalized the counts using the cpm function (log = TRUE) from the edgeR package v4.0.0^49^. The logFC (log Fold Change) was used as the slope value.

For GSEA, we analyzed Reactome pathways. We loaded pathways using the msigdbr function (species = “Homo sapiens”, category = “C2”, subcategory = “CP:REACTOME”) from the msigdbr package v7.5.1^50^. We ranked all genes, regardless of their adjusted P-values, by the following metrics:

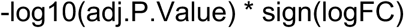

where adj.P.Value is the adjusted P value of the gene after Benjamini-Hochberg multiple comparison correction, logFC is the estimate of the log2-fold-change, and sign is the sign function. The ranked genes were used as input for the fgseaMultilevel function (minSize = 15, maxSize = 500, nproc = 1) from the fgsea package v1.28.0^51^ to run pre-ranked gene set enrichment analysis. For our pathway plot, we selected 11 immune pathways of interest from the list of Reactome pathways. The directionality of the enrichment score is based on the up- or down-regulation of gene expression rather than the absolute directionality of the signaling cascade.

### Sensitivity analysis of female SLE patients

For the female-stratified analysis, we analyzed the 240 bulk RNA-seq samples from CLUES participants who were female (sex assigned at birth). We normalized the gene counts using the voom function (normalize.method = “quantile”) from the limma package and fitted a linear model for the gene expression with the lmFit function using the follow design formula:

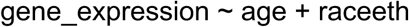

where gene_expression is the normalized expression of a given gene, age is the age of a sample’s subject, and raceeth is a categorical variable with four levels (White, Hispanic, Black, or Asian). Differential expression and GSEA were carried out as described for our primary analysis.

### Sensitivity analysis adjusting for corticosteroid treatment

For the steroid-adjusted analysis, we analyzed 267 bulk RNA-seq samples from CLUES participants with steroid dosage information in the corresponding metadata. We focused on patients who received moderate to high dose steroids, defined in our analysis as receipt of ≥ 7.5 mg of prednisone daily.

We normalized the gene counts using the voom function (normalize.method = “quantile”) from the limma package and fitted a linear model for the gene expression with the lmFit function using the following R design formula:

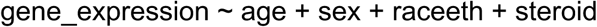

where gene_expression is the normalized expression of a given gene, age is the age of a sample’s subject, sex is a categorical variable with two levels and raceeth is a categorical variable with four levels as described above, and steroid is a factor with two levels (no steroid or yes steroid). Differential expression and gene set enrichment analyses were carried out as described for our primary analysis.

### Sensitivity analysis adjusting for hydroxychloroquine treatment

For the steroid-adjusted analysis, we analyzed 201 bulk RNA-seq samples from CLUES participants with Hydroxychloroquine dosage information in the corresponding metadata. We normalized the gene counts using the voom function (normalize.method = “quantile”) from the limma package and fitted a linear model for the gene expression with the lmFit function using the following R design formula:

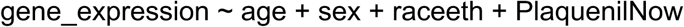

where gene_expression is the normalized expression of a given gene, age is the age of a sample’s individual, sex is a categorical variable with two levels and raceeth is a categorical variable with four levels as described above, and PlaquenilNow is a factor with two levels (no Hydroxychloroquine or yes Hydroxychloroquine). Differential expression and gene set enrichment analyses were carried out as described for our primary analysis.

### Sensitivity analysis adjusting for BMI

For the BMI-adjusted analysis, we analyzed all bulk RNA-seq samples from CLUES participants. We normalized the gene counts using the voom function (normalize.method = “quantile”) from the limma package and fitted a linear model for the gene expression with the lmFit function using the following R design formula:

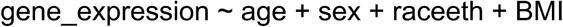

where gene_expression is the normalized expression of a given gene, age is the age of an individual, sex is a categorical variable with two levels and raceeth is a categorical variable with four levels as described above, and BMI is the BMI reading for the individual at the timepoint their sample was collected. Differential expression and gene set enrichment analyses were carried out as described for our primary analysis.

### Analysis of healthy control blood transcriptomic data from the Rotterdam Study

We analyzed normalized whole blood RNA microarray data from the Rotterdam Study, which is publicly available under Gene Expression Omnibus (GEO) accession GSE33828. As with SLE patients, we evaluated differential gene expression using linear modeling with age as a continuous variable, and controlled for sex (race/ethnicity information was not publicly available).

We analyzed all samples with > 10,000 protein coding genes that also had age and sex data available (n=880). To map probe IDs to gene symbols, we used the getGeo function (GEO = “GSE33828”) from the GEOquery package v2.70.0^52^. We used limma^47^ to analyze the Rotterdam Study microarray data which had already been normalized. We fitted a linear model for the gene expression with the lmFit function using the following R design formula:

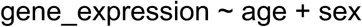

where gene_expression is the normalized expression of a given gene, age is the individual’s age, and sex is the individual’s sex. eBayes and topTable were used as described above. GSEA results were generated and analyzed similarly as described above.

For the slope scatter plot, the logFC values of the same leading-edge genes from the SLE analysis were used. The interferon score for each individual was calculated by computing the average normalized gene expression of the relevant genes.

### Interferon stimulated gene and interferon score analyses

Interferon subset analyses (slope scatter plot and IFN score) were conducted as follows. First, the leading-edge genes, or those which drive differential enrichment, were extracted from the IFN-α/β and IFN-γ pathways based on pathway results from the SLE data. The logFC values for these genes were used for the slope scatter plot.

For the interferon score analysis, the genes were split based on pathway. The interferon score was defined as the average normalized gene expression of these genes for each individual. The slope and P value for the effect of Age were extracted based on the following formula (variables described previously):

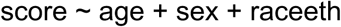

For the age-of-diagnosis analysis (AGEDX), we identified SLE samples with available diagnosis age information (n = 222). We treated AGEDX as a factor in our analysis (0 for AGEDX <= 50, 1 for AGEDX > 50). The slope and P value for the effect of age were extracted based on the following formula (variables described previously):

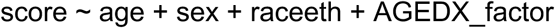

where AGEDX_factor is a categorical variable as described above, age is the individual’s age, sex is the individual’s sex, and raceeth is a categorical variable with four levels as described above.

### Proteomic analysis of SLE patients from the CLUES cohort

We measured the concentrations of 48 inflammatory proteins from 1 μL of plasma using the Olink proximity extension assay ‘Immune Surveillance’ panel (Olink Proteomics AB, Uppsala, Sweden). Proteins were measured from all SLE patients with available plasma (n = 268) following manufacturer’s instructions. Briefly, samples were incubated with oligonucleotide-labeled antibodies complementary to each protein for 18 h at 4°C. In the assay, partner probes are brought together in close proximity if a target protein is present, allowing the formation of a double-stranded oligonucleotide that can be quantified using a microfluidic real-time PCR instrument (Biomark HD, Fluidigm).

For each protein, we analyzed the relationship between its concentration (pg/mL) and age. For protein concentrations, any occurrences of NaN values were imputed as 0 to ensure the completeness of the dataset for statistical modeling. Additionally, any concentrations that did not meet QC standards (values such as “No Data” and “> ULOQ”) were removed from the analysis. Protein concentrations were log base 10 transformed with a small constant (1e-6). We fit a linear regression model for each protein concentration with the lm function using the following R design formula:

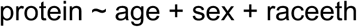

where protein is the concentration of that protein for a given sample, age is the age of a sample’s individual, sex is a categorical variable with two levels and raceeth is a categorical variable with four levels as described above. For our regression plot and volcano plots, we extracted the slope and P value for age. All P values were corrected using Benjamini-Hochberg multiple comparison correction.

### PBMC scRNA-seq analysis

We leveraged PBMC scRNA-seq data generated from 162 SLE cases and 48 healthy controls in the CLUES cohort^19^. The data was obtained from a publicly available dataset consisting of scRNA-seq of 1.2 million PBMCs from adult lupus and healthy control samples (https://cellxgene.cziscience.com/collections/436154da-bcf1-4130-9c8b-120ff9a888f2). There were 162 SLE cases from the CLUES cohort in this dataset, and 48 healthy controls from the UCSF Rheumatology Clinic.

The cell type annotation was performed as recently described^19^, and included two annotation levels: the first consisted of 11 “broad” cell types based on preliminary Louvain clustering, and the second of 14 lymphoid-specific cell subpopulations. The 11 broad cell types included CD14+ classical and CD16+ nonclassical monocytes (cM and ncM); conventional and plasmacytoid dendritic cells (cDC and pDC); CD4+ and CD8+ T cells (CD4 and CD8); natural killer cells (NK); B cells (B); plasmablasts (PB); proliferating T and NK cells (Prolif); and progenitor cells (Progen). The 14 lymphoid subpopulations included naïve, effector memory, and regulatory CD4+ T cells (CD4^+^ T_naïve_, CD4^+^ T_EM_, CD4^+^ T_reg_), naïve, GZMH+ cytotoxic, GZMK+ cytotoxic, and mucosal-associated invariant CD8+ T cells (CD8^+^ T_naïve_, CD8^+^ T_GZMH_, CD8^+^ T_GZMK_, T_MAIT_); CD56-bright and CD56-dim natural killer cells (NK_bright_, NK_dim_); naïve, memory, plasma, and atypical B cells (B_naïve_, B_mem_, B_plasma_, B_atypical_); and CD34+ progenitors (Progen). PBMC data was analyzed using Scanpy v1.9.3 and R v4.3.1, and visualized on a Uniform Manifold Approximation and Projection plot. For all analyses, we used linear modeling with age as a continuous variable and controlled for sex and race/ethnicity.

### Analysis of age-related changes in immune cell proportions from PBMC scRNA-seq data

From both SLE (n=148) and control patients (n=48) in the CLUES cohort, we created a multiple regression model for each of the 14 annotated cell types in the scRNA-seq data to analyze the relationship between age and cell proportion. To generate our multiple regression model, we used Python with the pandas package v1.2.4^53^ and the statsmodels package v0.12.2^54^. We used the ols function with the following design formula (in R notation):

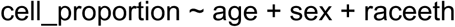

where cell_proportion is the proportion of a given cell type in the sample, age is the age of a sample’s individual, sex is a factor with two levels as described above, and raceeth is a categorical variable with two levels (European or Asian). To fit our model, we used the fit function (default settings) from the scikit-learn package v0.24.1^55^. We extracted the slope and associated P value from each model, and then adjusted the P value with Benjamini-Hochberg multiple comparison correction using the multipletests function from the statsmodels package.

### Differential gene expression analyses from PBMC scRNA-seq data

We used a pseudobulk approach for scRNA-seq differential gene expression analyses to avoid confounding from disproportionate cell type contributions from individual patients. This involved aggregating gene expression data for each cell type per patient. For each cell type, we retained samples with > 50 cells and > 5,000 protein-coding genes. For all groups and across the cell types, we retained protein-coding genes that had a minimum of 5 counts in at least 15% of the samples. Because too few pDC and Prolif cells were recovered, these cell types were not included in the differential expression analyses.

We normalized the gene counts using the voom function (normalize.method = “quantile”) from the limma package and fitted a linear model for the gene expression with the lmFit function using the follow R design formula:

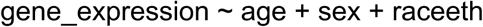

where gene_expression is the normalized expression of a given gene in the specific cell type, age is the age of a sample’s individual, sex is a categorical variable with two levels and raceeth is a categorical variable with two levels as described above. eBayes and topTable were used as described previously. GSEA results were generated and analyzed as described above in the bulk RNA-seq methods.

For the dot plots, we focused on four immune pathways found to differ significantly between groups in the bulk RNA-seq analysis. We used the logFC value from the topTable output as the slope. All P values were corrected using Benjamini-Hochberg multiple comparison correction.

For the SLE versus control sensitivity analysis in females at different age cutoffs, we aggregated gene expression data across all cell types, retaining samples with greater than 50 cells. We performed a similar quality control approach as for our bulk RNA-seq analysis, removing samples with fewer than 10,000 protein-coding genes and keeping genes with at least 10 counts in at least 20 percent of the samples. We split our samples into three groups based on the individual’s age at time of sample collection: <50, >50, and >65 years old. For each age group, we normalized the gene counts as described previously. We fit a linear model for gene expression with the lmFit function using the following R design formula:

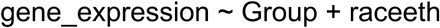

where gene_expression is the normalized expression of a given gene in the sample, raceeth is a factor with two levels, and Group is a factor with two levels, indicating whether the individual had SLE or was a control. eBayes and topTable were used as described previously. GSEA results were generated and analyzed as described previously.

### DNA methylation analysis

Methylation of genomic DNA was pre-processed using a previously detailed workflow^13^. Genomic DNA methylation from whole blood samples was processed using the Illumina Methylation EPIC BeadChip kit and R minfi package. Signal intensities were background subtracted and quantile normalized. After quality control steps, 723,424 CpG sites from 341 samples remained for analysis. Additionally, in this pre-processing stage, we filtered out probes that map to non-coding SNPs (61,037). Following this, we screened for sites mapping to non-autosomal chromosomes (15,833 CpG sites). Furthermore, patient filtering was conducted, initially excluding 13 patients with duplicated samples and those with incomplete information (2 patients). Finally, we filtered out patients who did not match the bulk RNA seq data (61 patients), resulting in a total of 646,554 CpG sites from 267 samples.

We employed the M values for the differential methylation analysis, specifically focusing on identifying individual probes associated with age. Utilizing limma v3.48.3, we implemented the Linear Model for Series of Arrays (lmfit), incorporating batch, race, and plate as covariates. To address multiple testing, we applied an FDR procedure using the Benjamini–Hochberg (BH) method, with significance defined at P < 0.05.

For classification, hypermethylated genes were defined as having a LogFC > 0, while hypomethylated genes had a LogFC < 0. Proximity of a CpG probe to a gene was determined using the Annotation for Illumina’s EPIC methylation arrays.

To explore overlaps between methylation at individual probes and differentially expressed genes in RNA-seq data, we employed a hypergeometric test in R’s phyper. Specifically, we compared hypermethylated CpGs (CpGs upregulated in relation to age) with negatively regulated RNA expression (transcriptome downregulated with age), and vice versa. Furthermore, we assessed Pearson correlations between RNA log fold change and methylation log fold change using sm_statCorr (smplot2 v 0.1.0), as well as correlations between methylation level and age for selected genes.

To analyze the relationship between methylation and gene expression, we performed Pearson correlation analysis between hypermethylated CpGs (n = 230) and corresponding genes with negatively regulated RNA expression with age (n = 91). The M value for each gene was computed by taking the average of the CpG data for that gene. P values for Pearson correlation were adjusted using Benjamini-Hochberg multiple correction.

Gene ontology enrichment analysis was conducted using the missMethyl package (v1.26.1), specifically employing the gometh function. We tested hypermethylated and hypomethylated regions independently, utilizing standard parameters. Pathways associated with the term “interferon” were selected.

Differentially Methylated Regions (DMRs) were investigated using the DMRcate package using lambda equal 100 and a scaling factor of 2. Results were corrected for multiple comparisons using BH.

## Data Availability

Genecounts derived from bulk RNA-seq data in the CLUES cohort are available from Gene Expression Omnibus (GEO) under accession number GSE277909. Genecounts from the Rotterdam Study control group are available under GEO accession GSE33828. scRNA-seq genecounts and fastq files are available under GEO accession GSE174188. Annotated scRNA-seq data are available at: https://cellxgene.cziscience.com/collections/436154da-bcf1-4130-9c8b-120ff9a888f2. Source data are provided in the source data file accompanying this manuscript. All code is available via Github at: https://github.com/infectiousdisease-langelier-lab/CLUES_aging.

## Supplementary Tables

**Table 1a.** Characteristics of Patients with SLE in Bulk RNA-seq Analyses.

| Characteristics | (n=271) |
| --- | --- |
| <u>Sociodemographic Factors:</u> |  |
| Age, mean ± SD | 44.5 ± 14.0 |
| Female | 240 (88.6%) |
| Race/Ethnicity |  |
| Asian | 98 (36.2%) |
| White | 82 (30.3%) |
| Hispanic | 64 (23.6%) |
| Black | 27 (10%) |
| <u>SLE Specific Characteristics:</u> |  |
| SLE disease duration, years, mean ± SD | 15.0 ± 9.8 |
| Disease activity by SLEDAI, mean ± SD | 2.9 ± 2.9 |
| Disease damage by SLICC, mean ± SD | 1.7 ± 1.9 |
| Lupus Severity Index, mean ± SD | 7.1 ± 1.6 |
| History of Lupus Nephritis | 115 (42.4) |
| Prednisone ≥ 7.5 mg/day | 56 (20.7%) |
| Current hydroxychloroquine use | 158 (58.3%) |
| <u>Comorbidities and Health Status:</u> |  |
| Cardiovascular Disease | 11 (4.1%) |
| Diabetes Mellitus, type 2 | 18 (6.6%) |
| History of malignancy | 13 (4.8%) |
| Obesity | 96 (35.4%) |
| Body Mass Index (kg/m<sup>2</sup>), mean ± SD | 26.2 ± 6.1 |
| Current nicotine use | 12 (4.4%) |

\*Values are percent unless otherwise indicated. P-values calculated using chi-squared tests for categorical measures and t-tests for continuous measures.
SLEDAI - Systemic Lupus Erythematosus Disease Activity Index
SLICC - Systemic Lupus International Collaborating Clinics Damage Index
CVD - history of stroke, coronary artery disease, and/or myocardial infarction
Obese defined as body mass index ≥ 30 kg/m<sup>2</sup>

**Table 1b.** Characteristics of Patients with SLE in scRNA-seq Analyses.

| Characteristics | (n=148) |
| --- | --- |
| <u>Sociodemographic Factors:</u> |  |
| Age, mean ± SD | 45.2 ± 13.8 |
| Female | 134 (90.5%) |
| Race/Ethnicity |  |
| Asian | 73 (49.3%) |
| White | 60 (40.5%) |
| Hispanic | 15 (10.1%) |
| Black | 0.0% |
| <u>SLE Specific Characteristics:</u> |  |
| SLE disease duration, years, mean ± SD | 16.5 ± 9.3 |
| Disease activity by SLEDAI, mean ± SD | 2.8 ± 2.7 |
| Disease damage by SLICC, mean ± SD | 1.5 ± 1.9 |
| Lupus Severity Index, mean ± SD | 6.8 ± 1.6 |
| History of Lupus Nephritis | 56 (37.8%) |
| Prednisone ≥ 7.5 mg/day | 29 (19.6%) |
| Current hydroxychloroquine use | 84 (56.8%) |
| <u>Comorbidities and Health Status:</u> |  |
| Cardiovascular Disease | 5 (3.4%) |
| Diabetes Mellitus, type 2 | 7 (4.7%) |
| History of malignancy | 9 (6.1%) |
| Obesity | 44 (29.7%) |
| Body Mass Index (kg/m<sup>2</sup>), mean ± SD | 25.1 ± 4.8 |
| Current nicotine use | 5 (3.4%) |

\*Values are percent unless otherwise indicated. P-values calculated using chi-squared tests for categorical measures and t-tests for continuous measures.
SLEDAI - Systemic Lupus Erythematosus Disease Activity Index
SLICC - Systemic Lupus International Collaborating Clinics Damage Index
CVD - history of stroke, coronary artery disease, and/or myocardial infarction
Obese defined as body mass index ≥ 30 kg/m<sup>2</sup>

**Table 1c.** Characteristics of Healthy Controls in scRNA-seq Analyses.

| Characteristics | (n=48) |
| --- | --- |
| <u>Sociodemographic Factors:</u> |  |
| Age, mean ± SD | 44.2 ± 16.8 |
| Female | 47 (97.9%) |
| Race/Ethnicity |  |
| Asian | 23 (47.9%) |
| White | 25 (52.1%) |
| Hispanic | 5 (10.4%) |
| Black | 0 (0.0%) |
| <u>Comorbidities and Health Status:</u> |  |
| Cardiovascular Disease<sup>1</sup> | 2 (4.2%) |
| Diabetes Mellitus, type 2<sup>2</sup> | 2 (4.8%) |
| History of malignancy<sup>3</sup> | 0 (0.0%) |
| Obesity<sup>4</sup> | 3 (7.0%) |
| Body Mass Index (kg/m<sup>2</sup>), mean ± SD<sup>4</sup> | 23.3 ± 3.7 |

\*Values are percent unless otherwise indicated. P-values calculated using chi-squared tests for categorical measures and t-tests for continuous measures. CVD = history of stroke, coronary artery disease, +/- myocardial infarction. Obesity defined as body mass index ≥ 30 kg/m<sup>2</sup>. <sup>1</sup>n=47, <sup>2</sup>n=42, <sup>3</sup>n=42, <sup>4</sup>n=43.

## Supplementary Figures

**Supp. Fig. 1.**
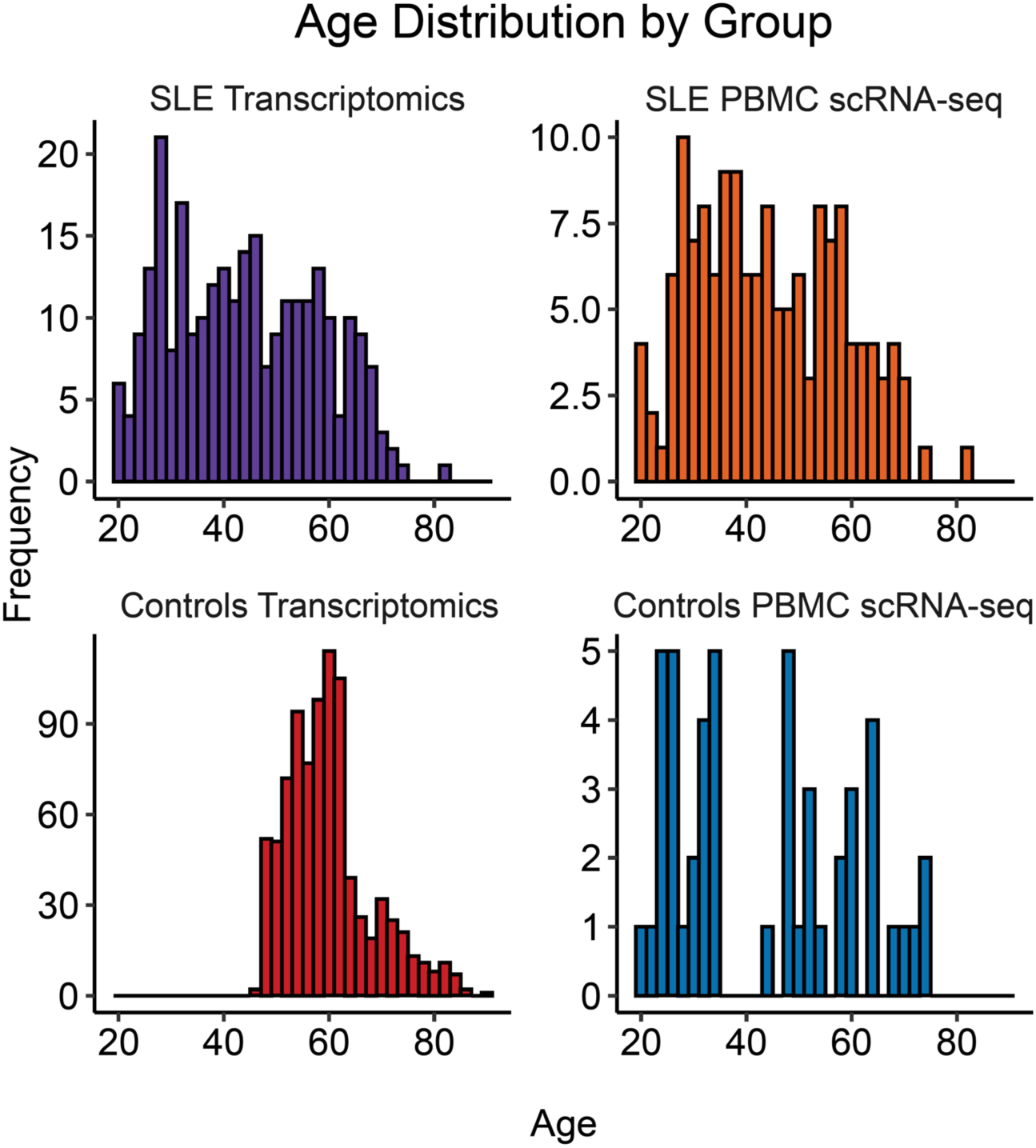
Age distribution of patients in the study cohorts. Histograms of ages of Patients with SLE in the CLUES cohort with bulk RNA-seq data (n=271), patients with SLE in the CLUES cohort with PBMC scRNA-seq data (n=148), healthy controls from the Rotterdam Study with whole blood RNA microarray data (n=880), and healthy controls from the CLUES cohort with PBMC scRNA-seq data (n=48).

**Supp. Fig. 2.**
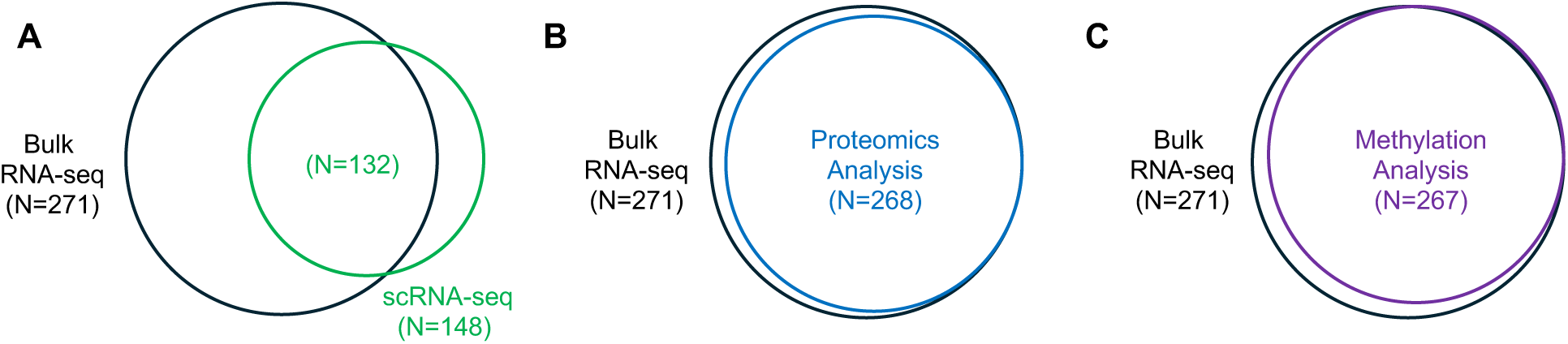
Overlap of analytic approaches for SLE patients in the CLUES cohort. **A** Overlap between bulk and scRNA-seq data from the 287 SLE patients studied in the CLUES cohort. **B** Of the 271 patients with bulk RNA-seq data, a subset also underwent proteomics analysis. **C** Of the 271 patients with bulk RNA-seq data, a subset also underwent methylation analysis.

**Supp. Fig. 3.**
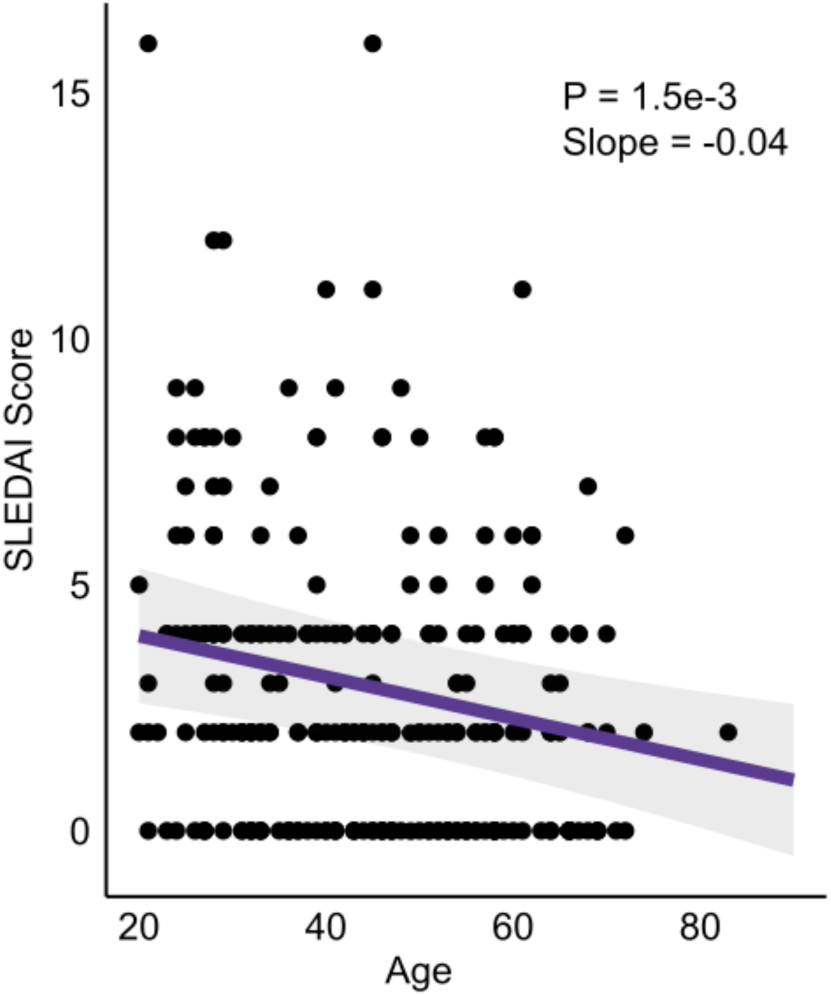
Relationship between Systemic Lupus Erythematosus Disease Activity Index (SLEDAI) score and age. n=271. Each data point refers to a patient’s SLEDAI score. The P-value was calculated with linear regression (solid purple line) based on the age coefficient. The ribbon indicates the 95% confidence interval of the linear regression fit.

**Supp. Fig. 4.**
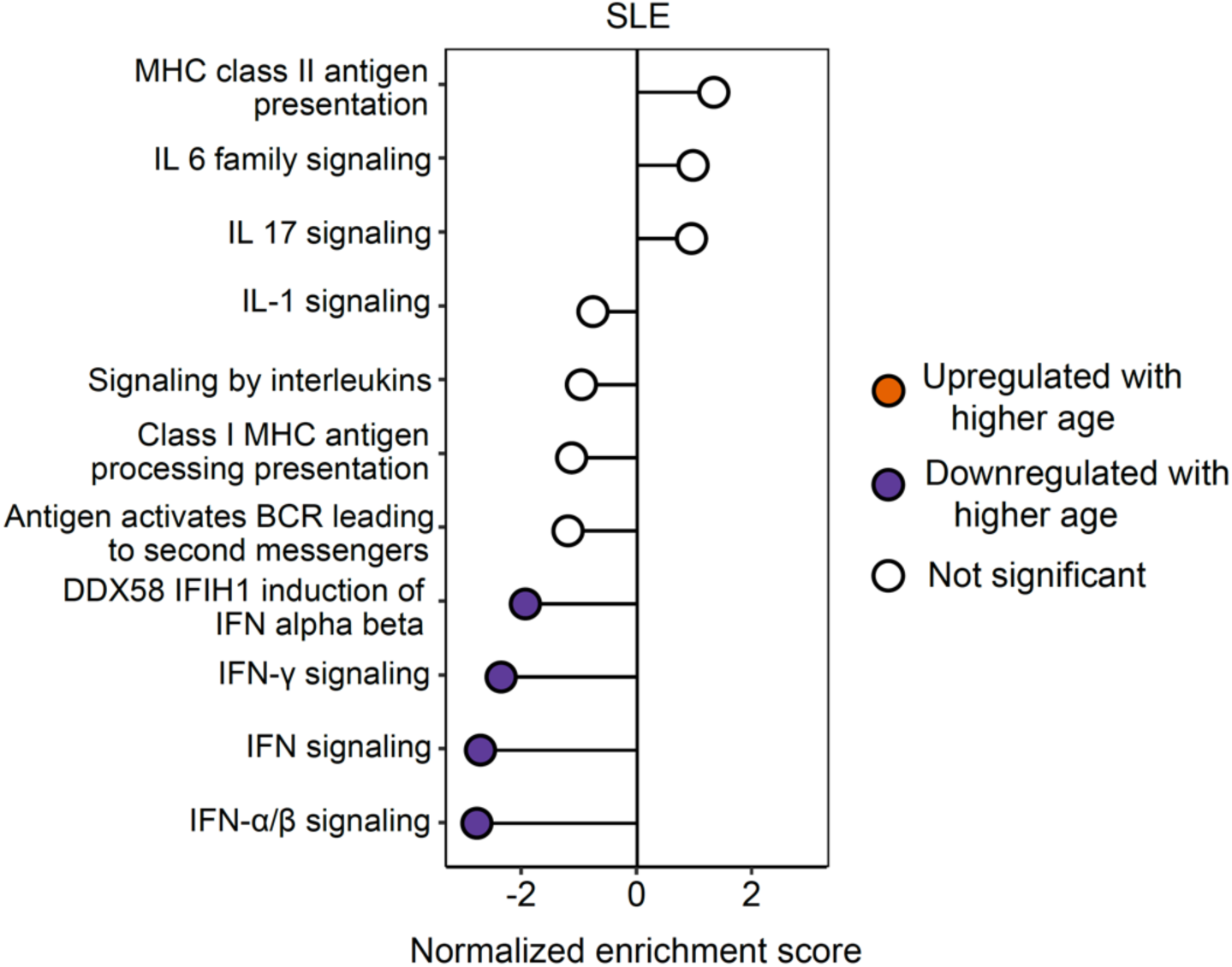
Sensitivity analysis in female SLE patients. GSEA results demonstrating immune signaling pathways associated with age in exclusively female SLE patients, adjusted for race/ethnicity (n=240). A positive normalized enrichment score value represents upregulation of the pathway with older age, and a negative value represents downregulation with older age. Filled circles represent pathways with a Benjamini-Hochberg adjusted P value < 0.05.

**Supp. Fig. 5.**
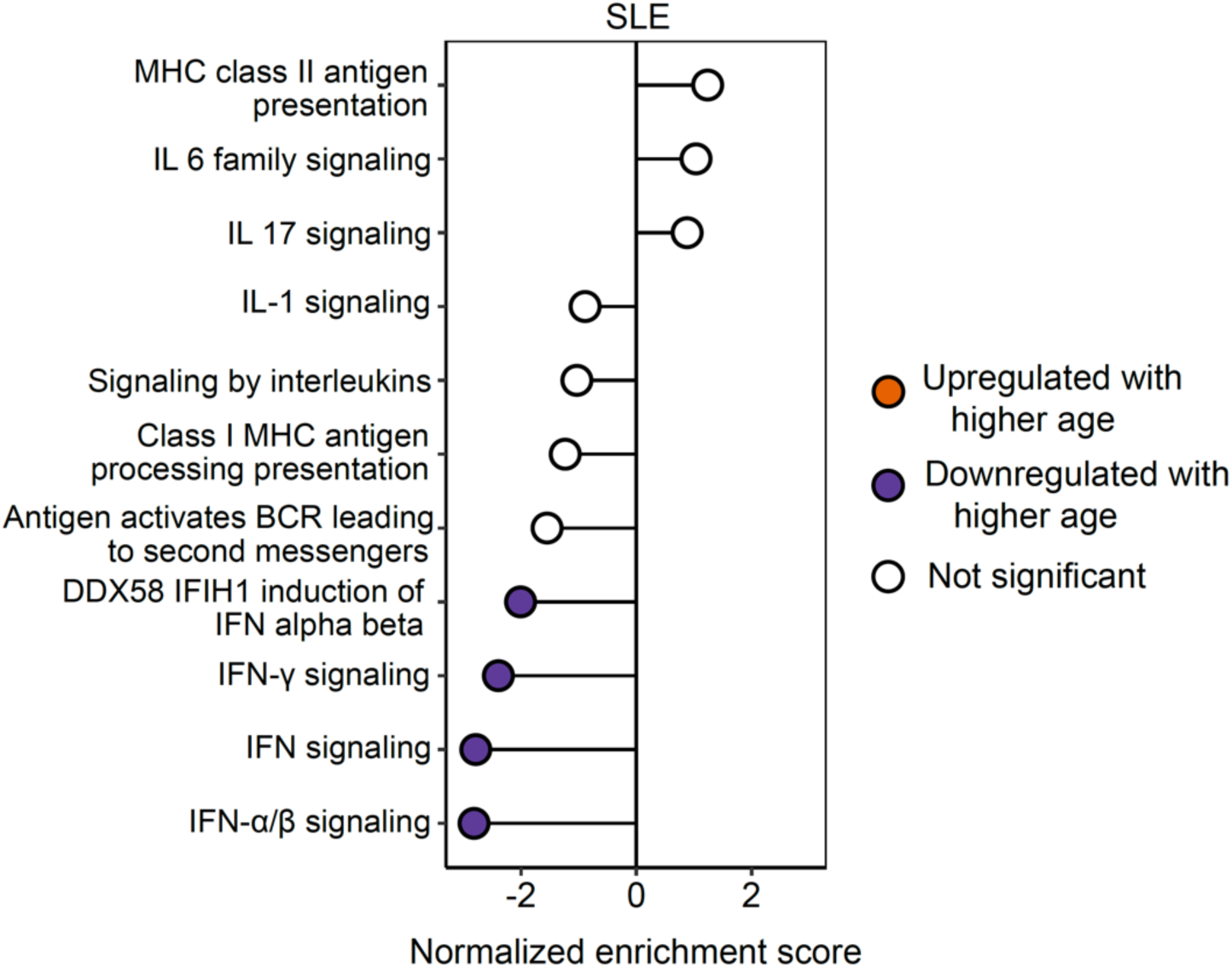
Sensitivity analysis adjusting for corticosteroid use. GSEA results demonstrating the immune signaling pathways in SLE patients associated with age, adjusted for sex, race/ethnicity and corticosteroid use (n=267). Corticosteroid use defined as > 7.5mg prednisone/day. A positive normalized enrichment score value represents upregulation of the pathway with older age, and a negative value represents downregulation with older age. Filled circles represent pathways with a Benjamini-Hochberg adjusted P value < 0.05.

**Supp. Fig. 6.**
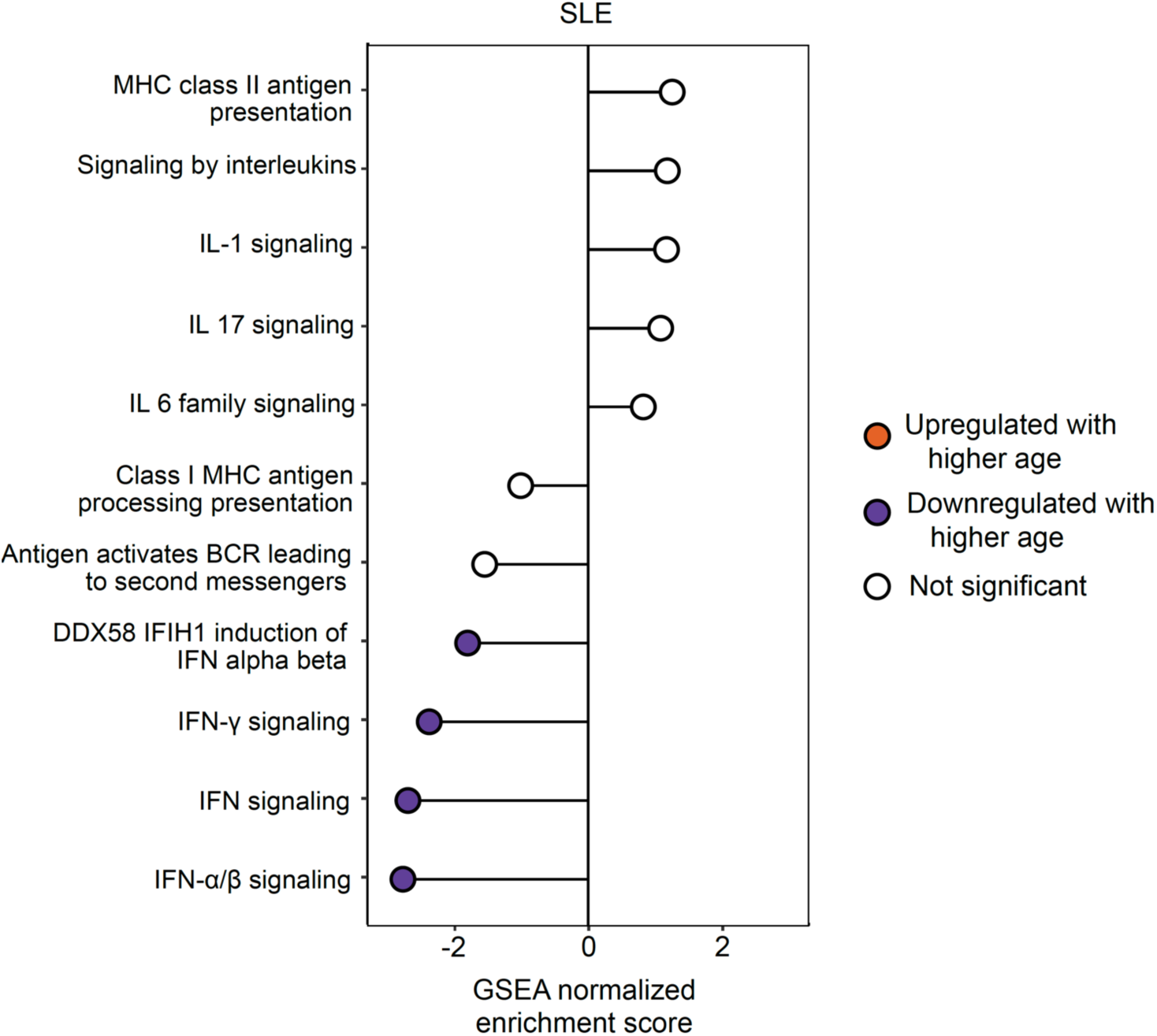
Sensitivity analysis adjusting for hydroxychloroquine use. GSEA results demonstrating the immune signaling pathways in SLE patients associated with age, adjusted for sex, race/ethnicity and hydroxychloroquine use (n=201). A positive normalized enrichment score value represents upregulation of the pathway with older age, and a negative value represents downregulation with older age. Filled circles represent pathways with a Benjamini-Hochberg adjusted P value < 0.05.

**Supp. Fig. 7.**
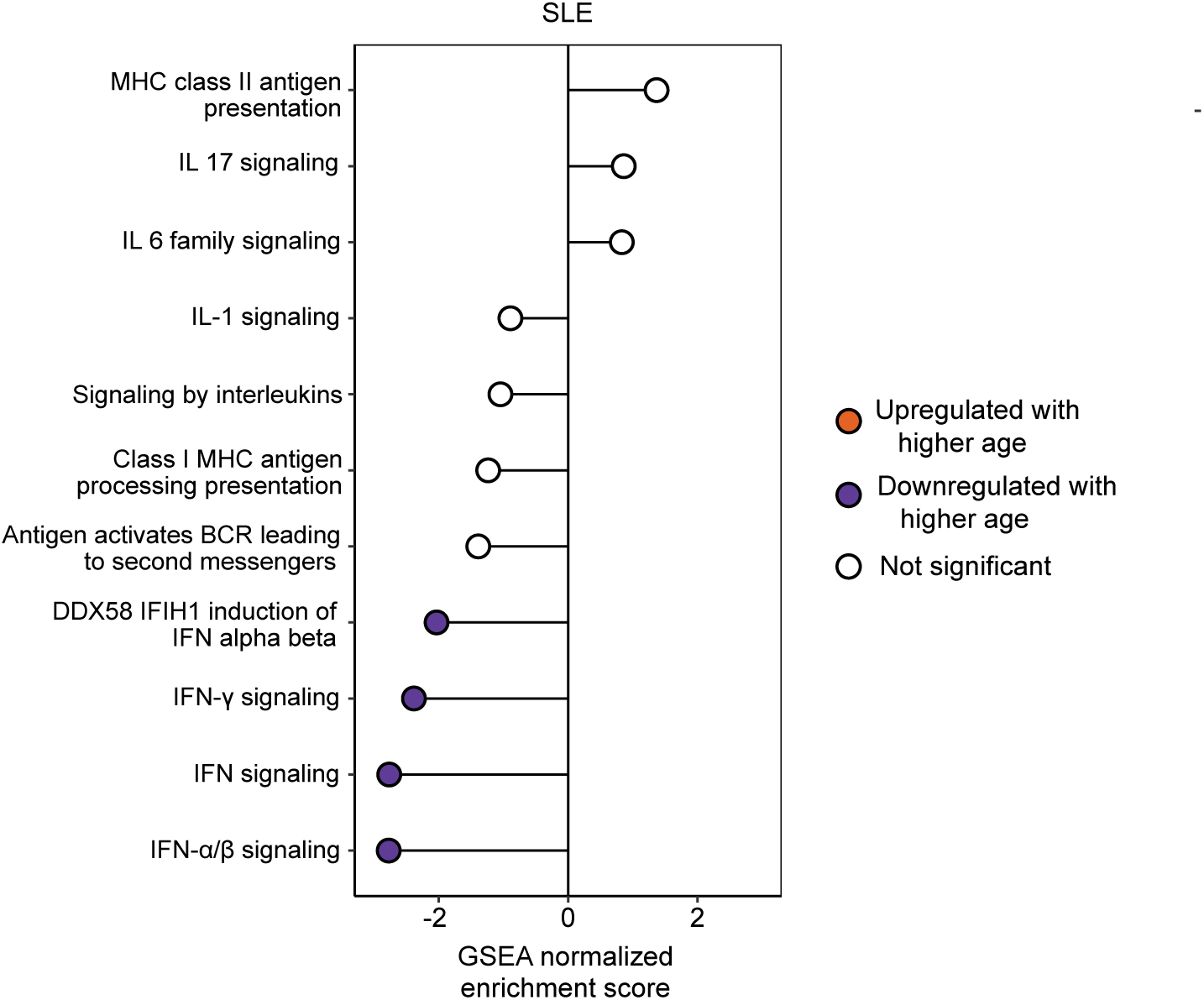
Sensitivity analysis adjusting for body mass index. GSEA results demonstrating the immune signaling pathways in SLE patients associated with age, adjusted for sex, race/ethnicity and body mass index (n=271). A positive normalized enrichment score value represents upregulation of the pathway with older age, and a negative value represents downregulation with older age. Filled circles represent pathways with a Benjamini-Hochberg adjusted P value < 0.05.

**Supp. Fig. 8.**
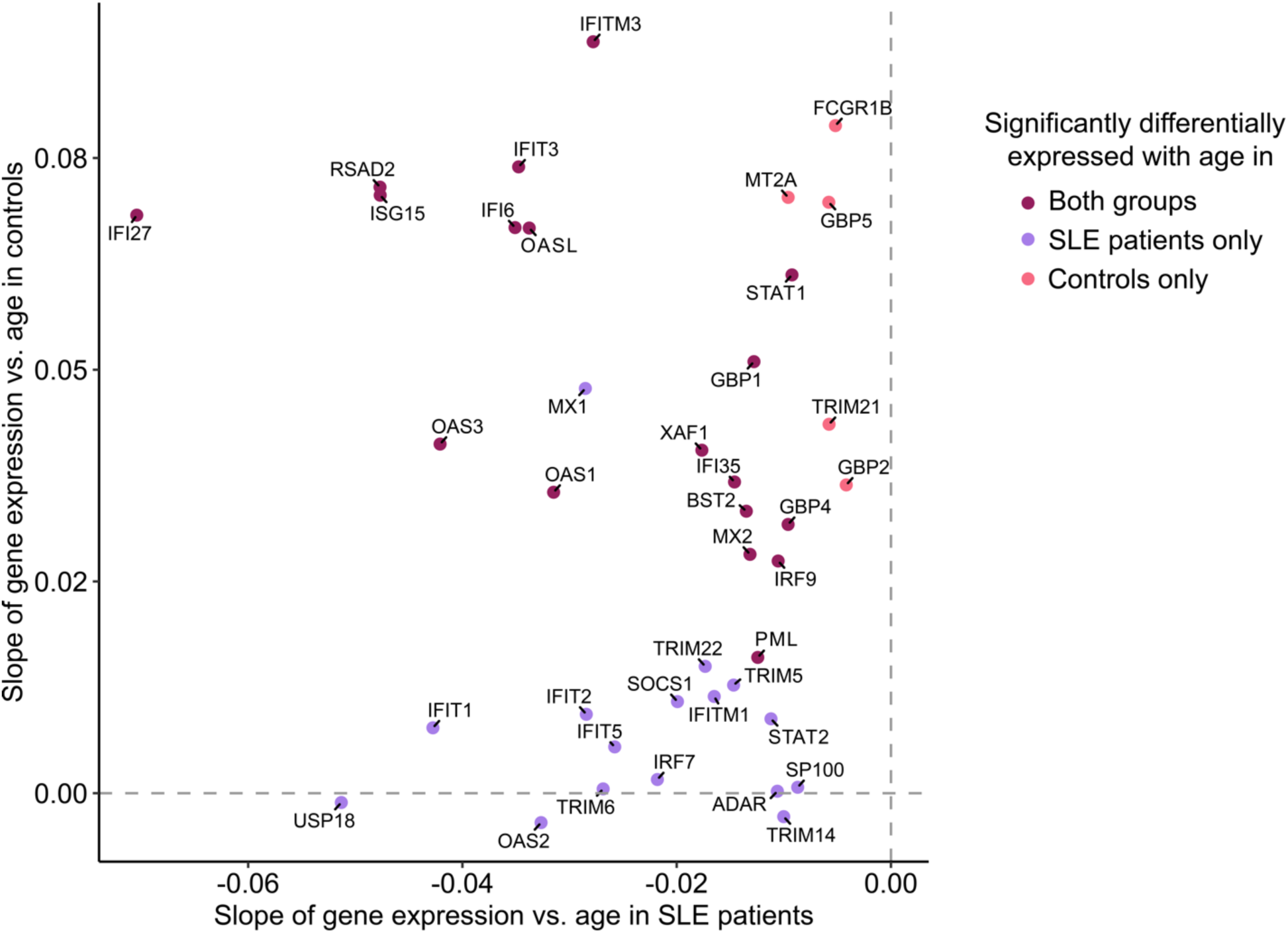
Interferon stimulated genes (ISGs) with age-related changes in expression. Scatter plot demonstrating slope of ISG expression versus age in SLE patients (X-axis) versus controls (Y-axis). ISGs with statistically significant age-related changes in expression in SLE patients only are shown in purple, controls only in light red, and both groups in dark red. The vertical dashed line indicates the position on the x-axis when the slope of gene expression vs. age in SLE patients is 0. The horizontal dashed line indicates the position on the y-axis when the slope of gene expression vs. age in SLE patients is 0.

**Supp. Fig. 9.**
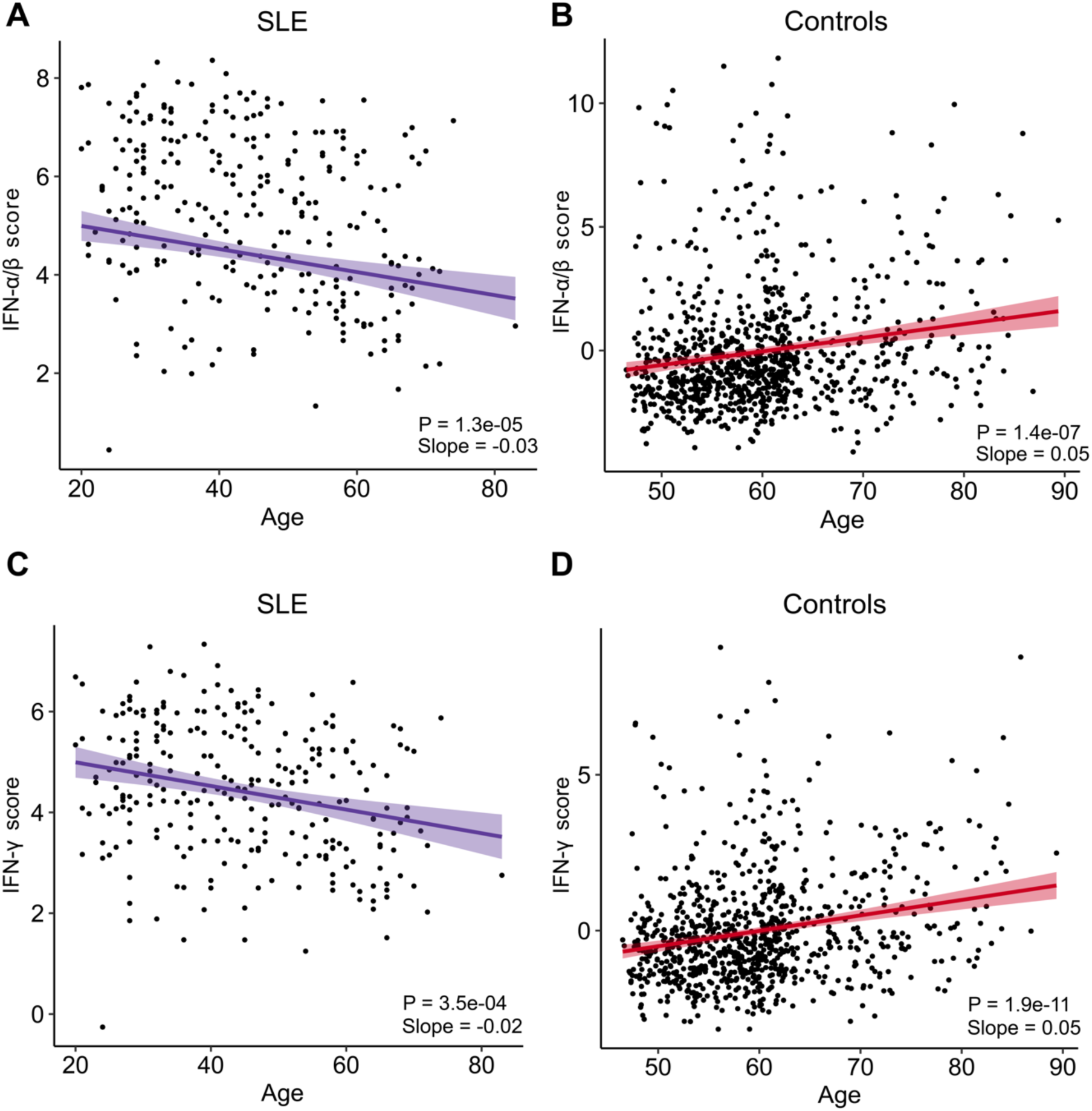
Relationship between interferon (IFN) scores and age for SLE patients and controls. **A** Scatter plot demonstrating average expression of leading-edge genes in the IFN-α/β signaling pathway (IFN-α/β score, Y-axis) versus age of SLE patients (X-axis). Slope and P value refer to the effect of age only. **B** Scatter plot demonstrating IFN-α/β score (Y-axis) versus age of controls (X-axis). C Scatter plot demonstrating average expression of leading-edge genes in the IFN-γ signaling pathway (IFN-γ score, Y-axis) versus age of SLE patients (X-axis). D Scatter plot demonstrating IFN-γ score (Y-axis) versus age of controls (X-axis).

**Supp. Fig. 10.**
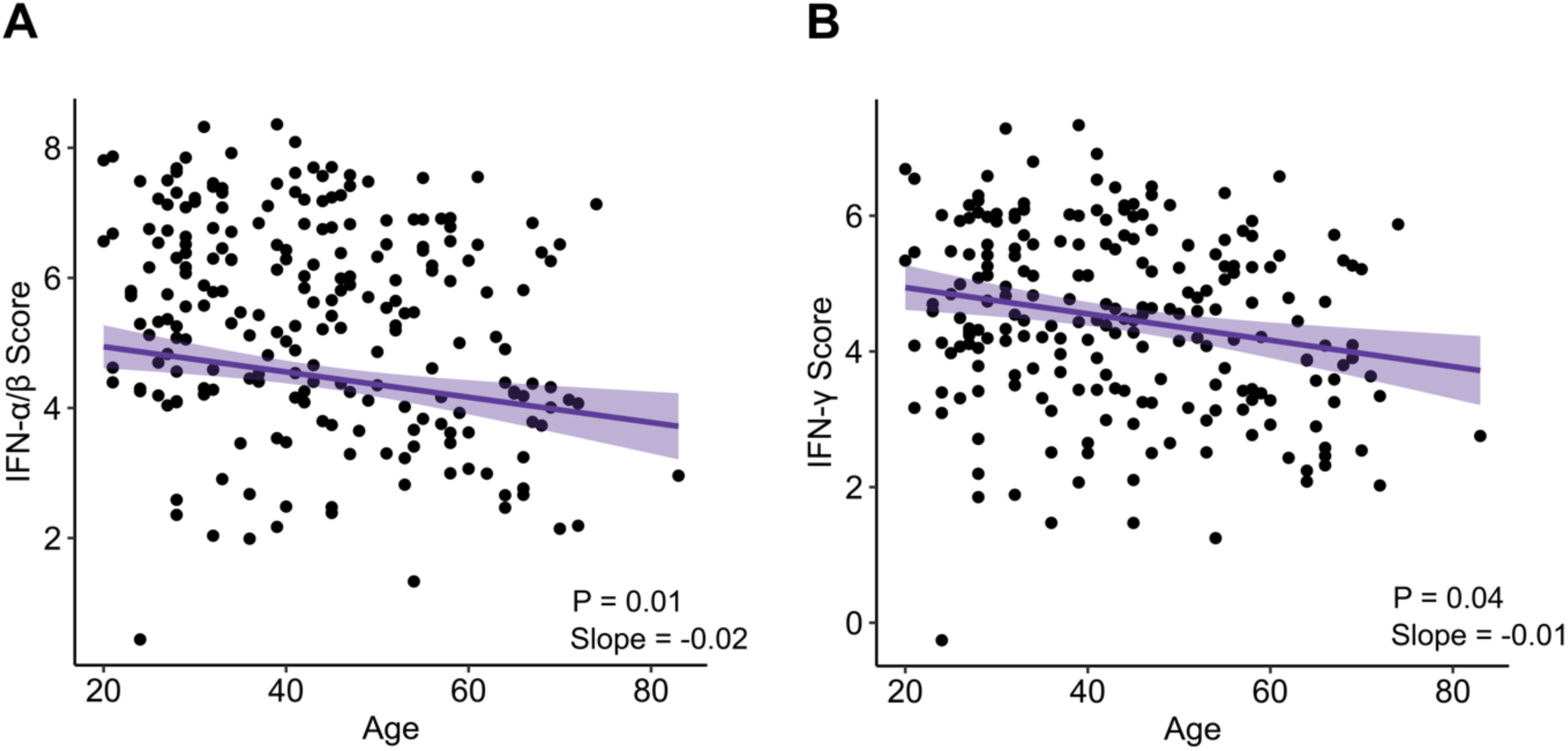
Relationship between interferon (IFN) scores and age for SLE patients controlling for age of SLE diagnosis. **A** Scatter plot demonstrating average expression of leading-edge genes in the IFN-α/β signaling pathway (IFN-α/β score, Y-axis) versus age of SLE patients (X-axis). Slope and P value refer to the effect of age only. **B** Scatter plot demonstrating IFN-α/β score (Y-axis) versus age of SLE patients (X-axis).

**Supp. Fig. 11.**
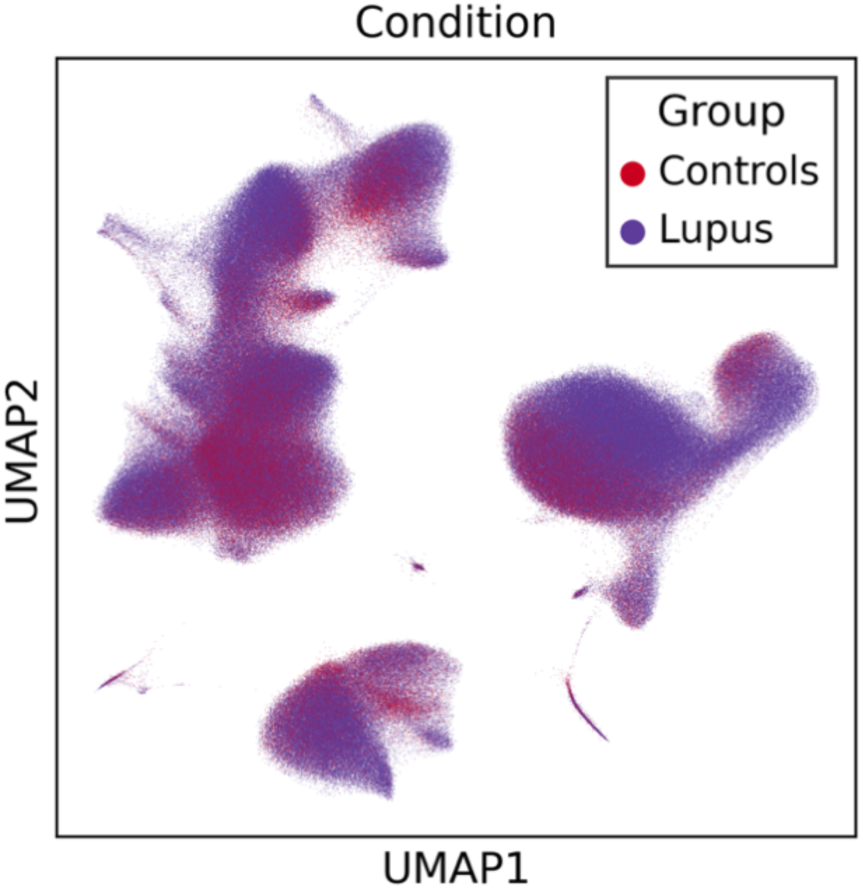
UMAP of immune cell populations based on SLE status. Uniform Manifold Approximation and Projection (UMAP) plot of scRNA-seq data from SLE patients in the CLUES cohort highlighting immune cell populations colored by SLE status. Purple dots correspond to single cells from SLE patients, (n=148 patients). Red dots correspond to single cells from control patients (n=48).

**Supp. Fig. 12.**
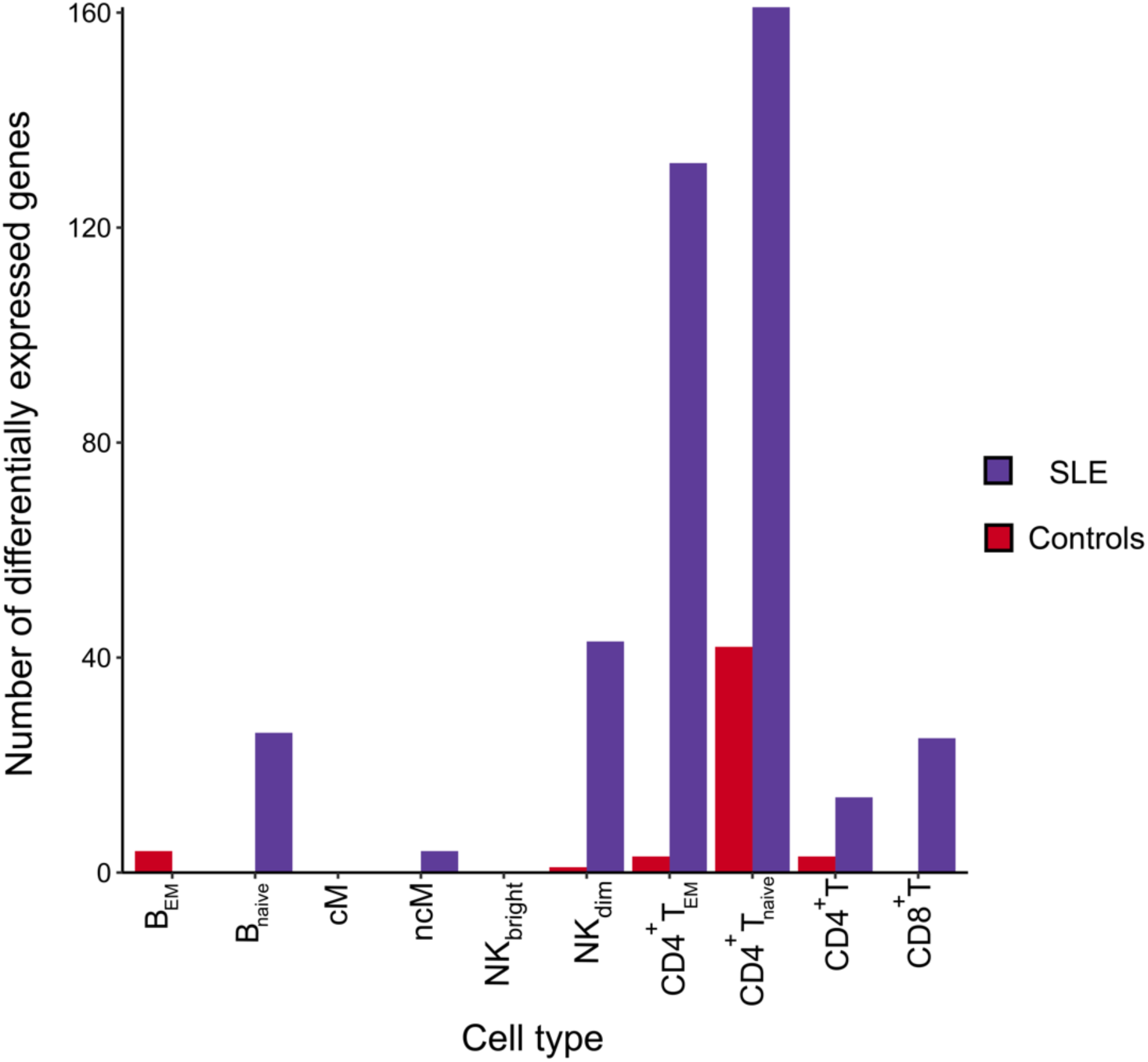
Differences in age-related changes in gene expression based on immune cell subpopulation. Grouped bar chart of the number of differentially expressed genes with age (P_adj_ < 0.05) by immune cell subpopulations for the pseudobulk analysis of PBMC scRNA-seq data. Purple refers to SLE patients (n=148) while red refers to control patients (n=48).

**Supp. Fig. 13.**
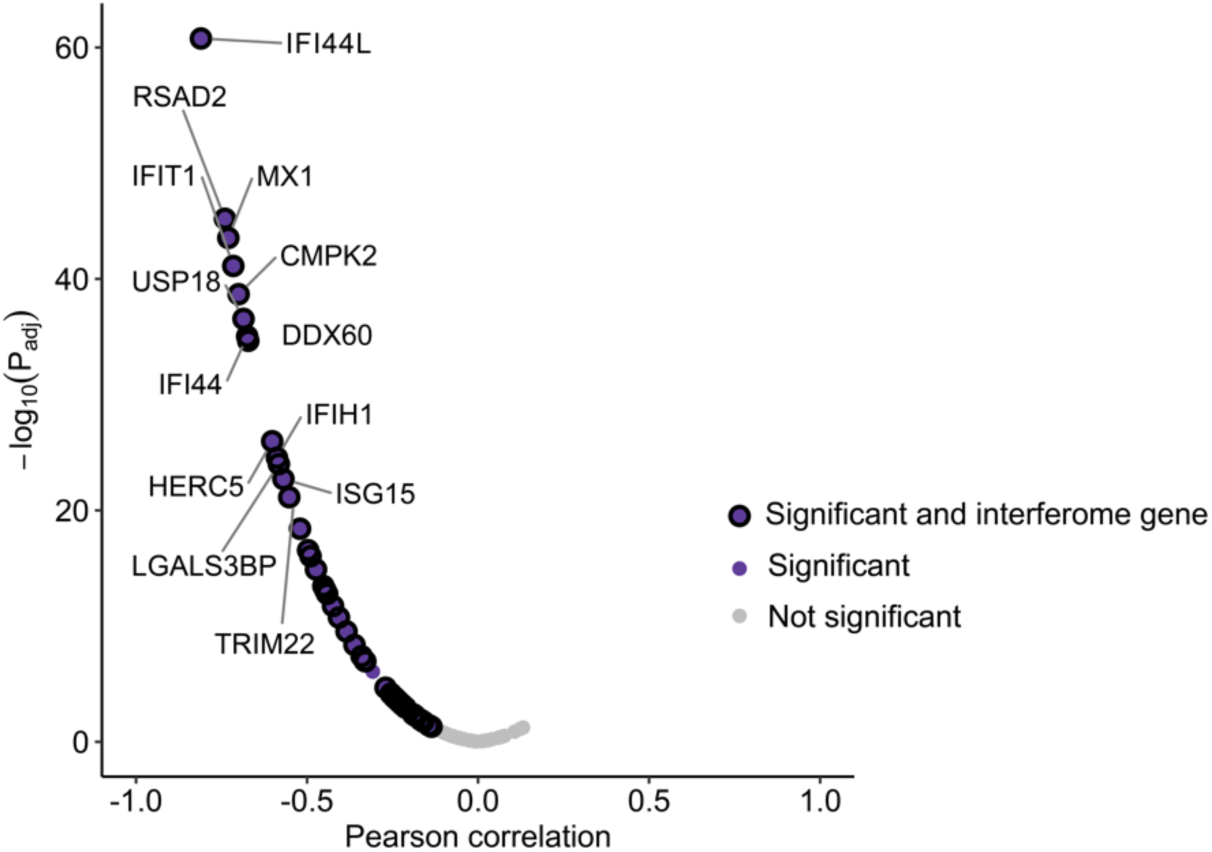
Correlation between methylation and normalized gene expression (downregulated genes, significantly hypermethylated CpGs). Volcano plot depicting the distribution of Pearson correlation coefficients between normalized gene expression values for genes significantly downregulated with age and M values for those genes based on the average of mapping hypermethylated CpGs. Purple dots represent coefficients with a Benjamini-Hochberg adjusted P value < 0.05. Larger dots with a thick, black outline represent coefficients for M and gene expression values for interferon-related genes. A negative Pearson correlation coefficient points to an inverse relationship between M value and gene expression; in particular, as the M value increases, gene expression decreases.

**Supp. Fig. 14.**
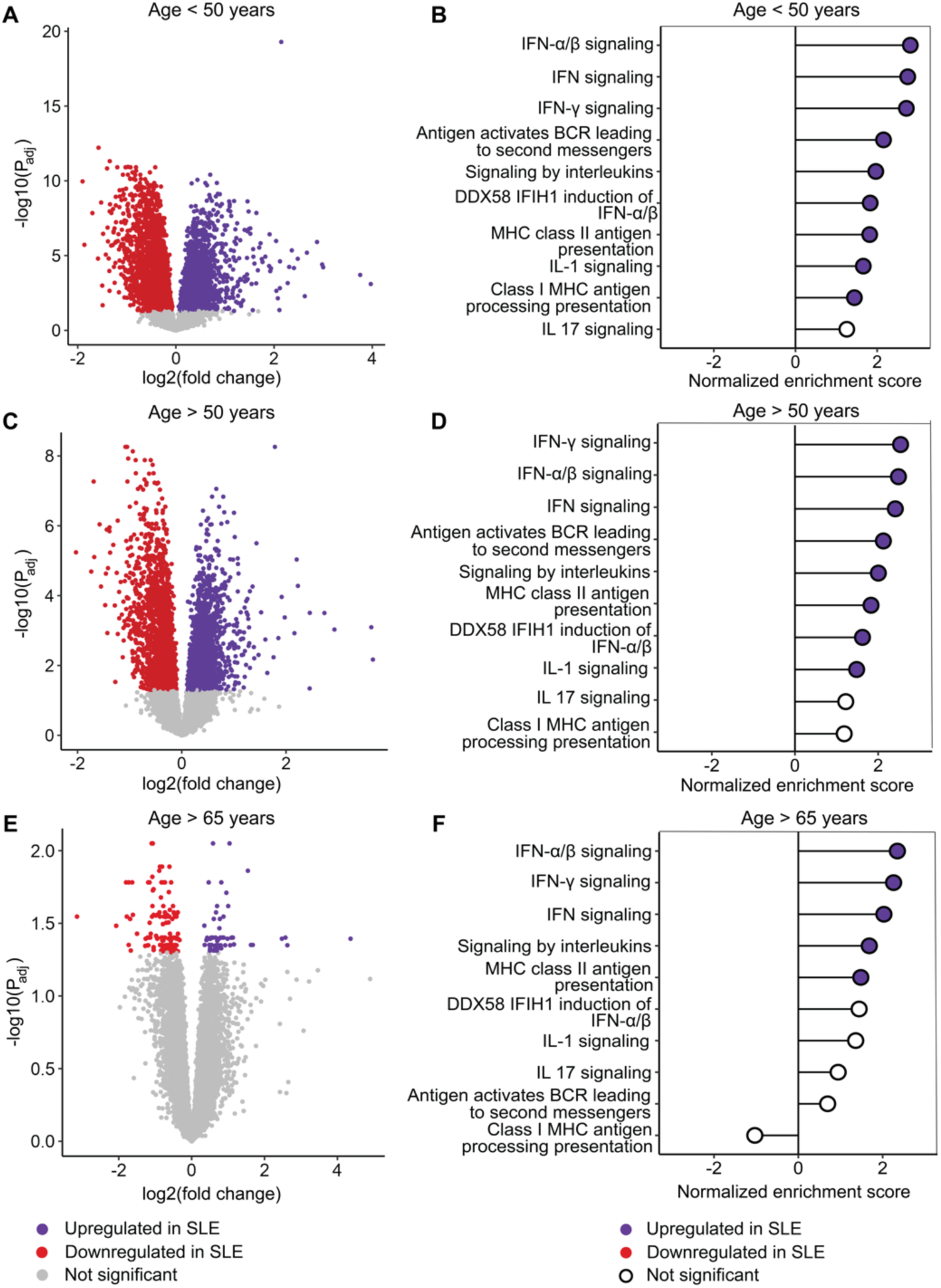
Differences in pseudobulk gene expression between female SLE patients and healthy female individuals, stratified by age group. **A** Volcano plot highlighting differentially expressed genes based on SLE status in female patients younger than 50 years old (n=110). Slope refers to the log2 fold change for each gene. **B** Gene Set Enrichment Analysis (GSEA) demonstrating immune-related biological signaling pathways differing based on SLE status in female patients younger than 50 years old. **C** Volcano plot highlighting differentially expressed genes based on SLE status in patients older than 50 years old (n=71). **D** GSEA demonstrating immune-related biological signaling pathways differing based on SLE status in patients older than 50 years old. **E** Volcano plot highlighting differentially expressed genes based on SLE status in patients older than 65 years old (n=17). **F** GSEA demonstrating immune-related biological signaling pathways differing based on SLE status in patients older than 65 years of age. Adjusted P values were calculated using the Benjamini-Hochberg method for all analyses.

## Supplementary Data Files

**Supp. Data 1.** Demographic data from Rotterdam Study healthy control cohort with bulk RNA-seq data.

**Supp. Data 2.** Demographic data from the CLUES healthy control cohort with scRNA-seq data.

**Supp. Data 3. A** Genes differentially expressed with age in SLE patients (n=271), adjusted for sex and race/ethnicity. **B** Gene set enrichment analysis (GSEA) of differentially expressed genes. Legend: logFC = log(2) fold change; padj = Benjamini-Hochberg adjusted P value; NES = normalized enrichment score; size = size of Reactome pathway; LeadingEdge = leading edge genes.

**Supp. Data 4. A** Genes differentially expressed with age in female SLE patients (n=240), adjusted for sex and race/ethnicity. **B** Gene set enrichment analysis (GSEA) of differentially expressed genes. Legend: logFC = log(2) fold change; padj = Benjamini-Hochberg adjusted P value; NES = normalized enrichment score; size = size of Reactome pathway; LeadingEdge = leading edge genes.

**Supp. Data 5. A** Genes differentially expressed with age in SLE patients, adjusted for receipt of corticosteroids, sex and race/ethnicity (n=267). **B** Gene set enrichment analysis (GSEA) of differentially expressed genes. Legend: logFC = log(2) fold change; padj = Benjamini-Hochberg adjusted P value; NES = normalized enrichment score; size = size of Reactome pathway; LeadingEdge = leading edge genes.

**Supp. Data 6. A** Genes differentially expressed with age in SLE patients, adjusted for receipt of hydroxychloroquine, sex and race/ethnicity (n=267). **B** Gene set enrichment analysis (GSEA) of differentially expressed genes. Legend: logFC = log(2) fold change; padj = Benjamini-Hochberg adjusted P value; NES = normalized enrichment score; size = size of Reactome pathway; LeadingEdge = leading edge genes.

**Supp. Data 7. A** Genes differentially expressed with age in SLE patients, adjusted for body mass index, sex and race/ethnicity (n=267). **B** Gene set enrichment analysis (GSEA) of differentially expressed genes. Legend: logFC = log(2) fold change; padj = Benjamini-Hochberg adjusted P value; NES = normalized enrichment score; size = size of Reactome pathway; LeadingEdge = leading edge genes.

**Supp. Data 8. A** Genes differentially expressed with age in healthy control patients, adjusted for sex and race/ethnicity (n=880). **B** Gene set enrichment analysis (GSEA) of differentially expressed genes. Legend: logFC = log(2) fold change; padj = Benjamini-Hochberg adjusted P value; NES = normalized enrichment score; size = size of Reactome pathway; LeadingEdge = leading edge genes.

**Supp. Data 9.** Linear regression results for protein concentrations and age in SLE patients from the CLUES cohort, adjusting for sex and race/ethnicity. Legend: Protein = protein; Slope = coefficient of age; Adjusted-P-Value = Benjamini-Hochberg adjusted P value, P = naïve p value, n = number of samples in analysis.

**Supp. Data 10.** Pearson correlation values between expression for genes (n = 91) downregulated with age and average M values calculated from hypermethylated mapping CpGs. Legend: Correlation = Pearson correlation; P_value = P value for Pearson correlation; Adj_P_value = Benjamini-Hochberg adjusted P value; Interferome = is the gene part of the interferome.

